# Functional investigation of inherited noncoding genetic variation impacting the pharmacogenomics of childhood acute lymphoblastic leukemia treatment

**DOI:** 10.1101/2023.02.10.23285762

**Authors:** Kashi Raj Bhattarai, Robert J. Mobley, Kelly R. Barnett, Daniel C. Ferguson, Baranda S. Hansen, Jonathan D. Diedrich, Brennan P. Bergeron, Wenjian Yang, Kristine R. Crews, Christopher S. Manring, Elias Jabbour, Elisabeth Paietta, Mark R. Litzow, Steven M. Kornblau, Wendy Stock, Hiroto Inaba, Sima Jeha, Ching-Hon Pui, Cheng Cheng, Shondra M. Pruett-Miller, Mary V. Relling, Jun J. Yang, William E. Evans, Daniel Savic

**Affiliations:** Hematological Malignancies Program, St. Jude Children’s Research Hospital, Memphis, TN; Department of Pharmacy and Pharmaceutical Sciences, St. Jude Children’s Research Hospital, Memphis, TN; Center for Advanced Genome Engineering, St. Jude Children’s Research Hospital, Memphis, TN 38105, USA; Department of Cell and Molecular Biology, St. Jude Children’s Research Hospital, Memphis, TN 38105, USA; Graduate School of Biomedical Sciences, St. Jude Children’s Research Hospital, Memphis, TN; Alliance Hematologic Malignancy Biorepository; Clara D. Bloomfield Center for Leukemia Outcomes Research, Columbus, OH 43210, USA; Department of Leukemia, The University of Texas MD Anderson Cancer Center, Houston, TX; Albert Einstein College of Medicine, New York, NY; Division of Hematology, Department of Medicine, Mayo Clinic, Rochester, MN 55905, USA; Comprehensive Cancer Center, University of Chicago Medicine, Chicago, IL; Department of Oncology, St. Jude Children’s Research Hospital, Memphis, TN; Department of Biostatistics, St. Jude Children’s Research Hospital, Memphis, TN; Integrated Biomedical Sciences Program, University of Tennessee Health Science Center, Memphis, TN

## Abstract

Although acute lymphoblastic leukemia (ALL) is the most common childhood cancer, there is limited understanding of the contribution of inherited genetic variation on inter-individual differences in chemotherapy response. Defining genetic factors impacting therapy failure can help better predict response and identify drug resistance mechanisms. We therefore mapped inherited noncoding variants associated with chemotherapeutic drug resistance and/or treatment outcome to ALL *cis*-regulatory elements and investigated their gene regulatory potential and genomic connectivity using massively parallel reporter assays and promoter capture Hi-C, respectively. We identified 53 variants with reproducible allele-specific effects on transcription and high-confidence gene targets. Subsequent functional interrogation of the top variant (rs1247117) determined that it disrupted a PU.1 consensus motif and PU.1 binding affinity. Importantly, deletion of the genomic interval containing rs1247117 sensitized ALL cells to vincristine. Together, these data demonstrate that noncoding regulatory variation associated with diverse pharmacological traits harbor significant effects on allele-specific transcriptional activity and impact sensitivity to chemotherapeutic agents in ALL.

## INTRODUCTION

Due to continual advances in treatment protocol over the last 60 years, the survival rate of the most common malignancy in children, acute lymphoblastic leukemia (ALL), has dramatically improved to over 90% in the high-income countries (*1*). Despite these advances, survival rates of pediatric patients experiencing refractory or relapsed ALL were only 30-50% and those of adults were especially low (∼10%) (*2*). Thus, improving the understanding of the underlying genetic risk factors impacting response to ALL chemotherapy is a major step in improving outcomes for patients with refractory or relapsed ALL.

Genome-wide association studies (GWAS) have identified numerous inherited DNA sequence variants associated with treatment outcome in childhood ALL from clinical trials carried out by St. Jude Children’s Research Hospital and the Children’s Oncology group (*3-5*). This includes GWAS analyses that identified inherited genetic contributors associated with patient relapse (*4, 5*) and persistence of minimal residual disease (MRD) after induction chemotherapy (*3*), which is an early indicator of treatment failure (*6-9*). In addition, *ex vivo* chemotherapeutic drug sensitivity testing using primary ALL cells from patients serves as an informative pharmacological phenotype (*10*). When integrated with genotype profiling for GWAS, these analyses identify variants contributing to antileukemic drug resistance that reflects *in vivo* and *ex vivo* resistance and is therefore predictive of treatment outcome in patients (*10-22*).

Because most GWAS variants, including pharmacogenomic variants (*23, 24*), lie in noncoding sequences in the human genome, their connection to gene regulation and cellular biology has yet to be established. Moreover, given that dozens of variants are typically in strong linkage disequilibrium (LD) with an associated sentinel variant, pinpointing causal variants at GWAS loci has been challenging. Noncoding GWAS variants have been consistently linked to disruption of *cis*-regulatory element (CRE) activity and gene regulation (*25*). As a result, the functional evaluation of these regulatory variants involves an examination of their allele-specific activities on transcriptional output which has traditionally been a low-throughput endeavor. Therefore, the functional investigation of all associated regulatory variation at GWAS loci (sentinel and LD) proved to be an intractable hurdle to investigators. Recent technological advances however have ameliorated these challenges through the advent of massively parallel reporter assays (MPRAs) where the reporter aspect is often a self-transcribed barcode in the 3’ UTR of a reporter gene that is detected using next-generation sequencing. MPRAs allow the simultaneous, rapid and robust detection of differences in transcriptional output from a library of *cis*-regulatory sequences of interest (*26-28*). MPRAs have since been applied to the study of regulatory variation at GWAS loci through an examination of allele-specific effects on reporter gene expression (*29-35*).

Another challenge is connecting regulatory variation at promoter-distal CREs to a target gene, as the closest gene may not be the target gene (*36*). To circumvent these challenges, regulatory variation can be coupled to transcriptomics to identify variants impacting the expression of a candidate target gene through expression quantitative trait locus (eQTL) mapping (*36, 37*). Functional genomics offer additional solutions through the mapping of three-dimensional (3D) genomic interactions (*38*). Because promoter-distal CREs (e.g., enhancers) regulate gene expression through long-range 3D looping to the promoters of target genes (*25*), an attractive assay to identify gene targets of promoter-distal regulatory variants is promoter capture Hi-C (promoter CHiC) (*39*). Promoter capture Hi-C and related 3D chromatin interaction assays have been implemented at multiple GWAS loci to identify gene targets of promoter-distal regulatory variation (*29, 36, 40-43*).

To better understand the underlying genetic and gene regulatory factors that impact diverse pharmacological traits in ALL, we performed a comprehensive functional interrogation of GWAS regulatory variants that map to ALL accessible chromatin sites and that are associated with *ex vivo* chemotherapeutic drug resistance in primary ALL cells from patients and/or ALL treatment outcome (i.e., relapse and persistence of MRD) in patients using MPRA. We coupled these results with promoter CHiC to identify candidate target genes of functional regulatory variants with significant allele-specific effects on reporter gene expression. Finally, we functionally investigated the impact of the top regulatory variant on transcription factor binding, neighboring gene expression and chemotherapeutic drug resistance in ALL cell lines. To our knowledge, this study represents the largest functional investigation of regulatory variants impacting the pharmacogenomics of chemotherapy treatment and fills an unmet need for large-scale functional examinations of regulatory GWAS variants associated with pharmacological traits.

## RESULTS

### Identification of regulatory variants impacting the pharmacogenomics of ALL treatment

Single nucleotide variants (SNVs) impacting diverse pharmacological traits in ALL were identified for functional interrogation. We chose SNVs associated with relapse or persistence of MRD after induction chemotherapy in childhood ALL patients to investigate the role of inherited regulatory variants impacting clinical phenotypes (i.e., treatment outcome). These SNVs were identified from published GWAS of ALL patients enrolled in St. Jude Children’s Research Hospital and the Children’s Oncology Group clinical protocols (*3-5*) (see **Methods** for SNV selection criteria). Variant selection included additional prioritization for SNVs associated with drug resistance phenotypes in primary ALL cells to enrich for variation impacting ALL cell biology. These treatment outcome-associated variants, as well as all variants in high LD (r^2^>0.8) with the sentinel GWAS variants, were further evaluated (**Fig 1A-B**).

**Figure 1:**
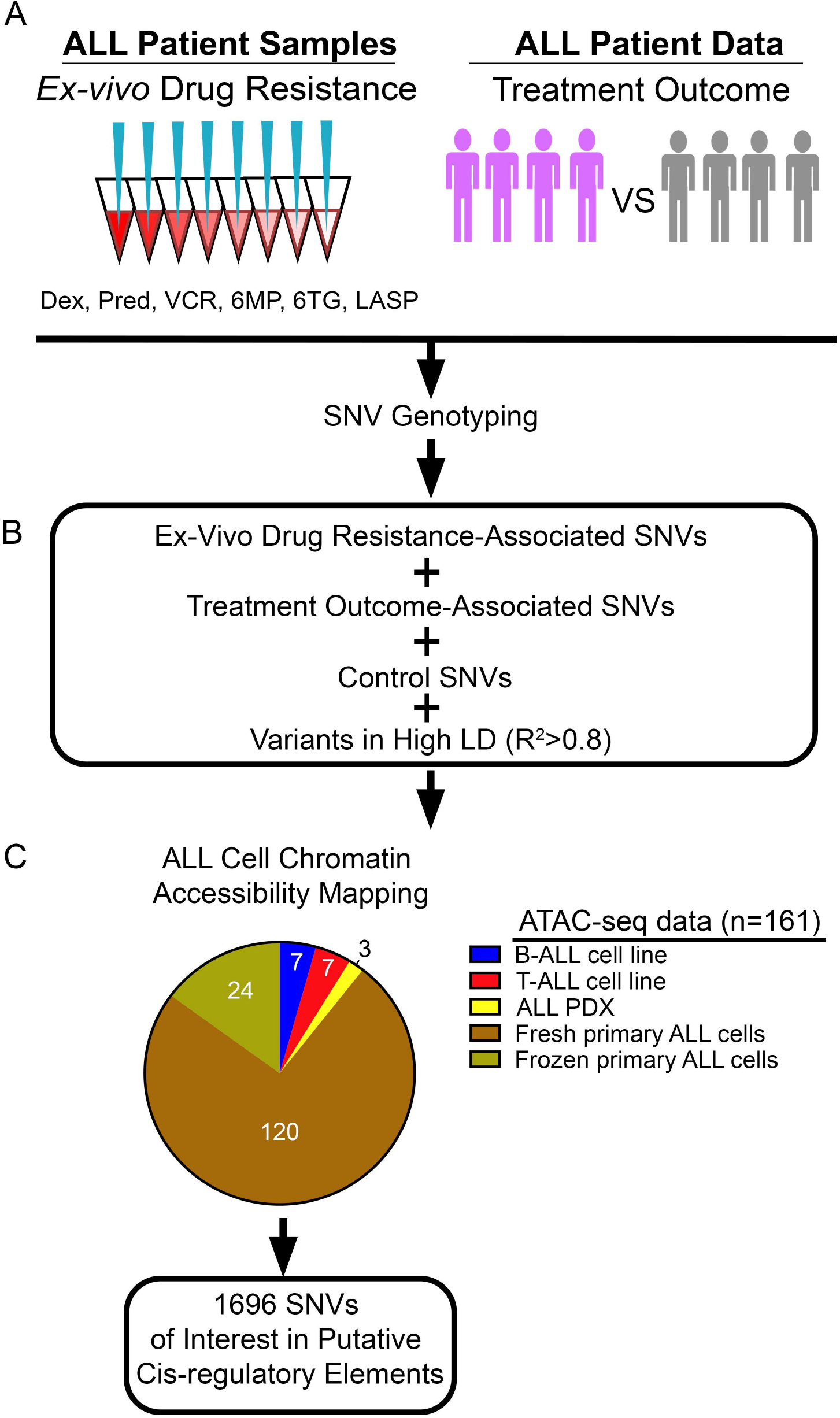
Identification and mapping of regulatory variants impacting the pharmacogenomics of ALL treatment. **(A)** SNVs of interest from GWAS were pursued based on association with *ex vivo* chemotherapeutic drug resistance in primary ALL cells from patients and/or treatment outcome. Dex= dexamethasone, Pred= prednisolone, VCR= vincristine, 6MP= 6-mercaptopurine, 6TG= 6-thioguanine, LASP= L-asparaginase. **(B)** GWAS SNVs were combined with ALL disease susceptibly control GWAS SNVs and SNVs in high LD (R^2^>0.8) and **(C)** mapped to accessible chromatin sites in ALL cell lines, ALL PDXs and primary ALL cells from patients.

We also identified variants directly associated with *ex vivo* chemotherapeutic drug resistance in primary ALL cells from patients by performing GWAS analyses using SNV genotype information and *ex vivo* drug resistance assay results for six antileukemic agents (prednisolone, dexamethasone, vincristine, L-asparaginase, 6-mercaptopurine and 6-thioguanine) in primary ALL cells from 312-344 patients enrolled in the Total Therapy XVI clinical protocol at St. Jude Children’s Research Hospital (see **Methods**). We further prioritized for functional *ex vivo* drug resistance SNVs by examining if they were eQTLs in primary ALL cells or in EBV-transformed lymphocytes from the Genotype-Tissue Expression (GTEx) consortium (*37*). All *ex vivo* drug resistance-associated eQTL variants, as well as variants in high LD (r^2^>0.8) with these sentinel GWAS variants, were further evaluated (**Fig 1A-B**).

GWAS have also been performed for childhood ALL disease susceptibility and identified several GWAS loci harboring variants with genome-wide significance (*44-50*). Several follow-up studies of these GWAS loci have identified candidate causal noncoding regulatory variants and mechanisms involving gene regulatory disruptions (*51-53*). As a result, we used ALL disease susceptibility variants, as well as variants in high LD (r^2^>0.8) with them, as positive controls in our study (**Fig 1A-B**).

Because most of these variants map to noncoding portions of the human genome, these data point to disruptions in gene regulation as the underlying mechanism of how these variants impact ALL cell biology. We therefore utilized assay for transposase-accessible chromatin with high-throughput sequencing (ATAC-seq) (*54*) chromatin accessibility data in 158 ALL cell models, comprised of primary ALL cells (cryopreserved, n=24 (*55*); fresh, n=120), ALL cell lines (n=14) and ALL patient-derived xenografts (PDXs, n=3), to uncover which variants map to putative CREs in ALL cells (*56*) (i.e., regulatory variants; **Fig 1C**). ATAC-seq data from primary ALL, ALL cell lines and PDXs were combined and identified 1696 regulatory variants at accessible chromatin sites in ALL cells for functional investigation (**Fig 1C** and **Sup File 1**).

### Assessing the impact of regulatory variation on transcriptional output using MPRA

To examine the functional effects of these 1696 regulatory variants on transcriptional output in a high-throughput manner we utilized a barcode-based MPRA platform (*29, 32*) to measure differences in allele-specific transcriptional output (**Fig 2A**). Oligonucleotides containing 175-bp of genomic sequence centered on each reference (ref) or alternative (alt) variant allele, a restriction site, and a unique 10-bp barcode sequence were cloned into plasmids. An open reading frame containing a minimal promoter driving GFP was then inserted at the restriction site between the alleles of interest and their unique barcodes (**Fig 2A**). We utilized 28 unique 3’UTR DNA barcodes per variant allele (56 barcodes per regulatory variant), and variants near bidirectional promoters (47 total variants) were tested using both sequence orientations. In total, 97,608 variant-harboring oligonucleotides were evaluated for allele-specific differences in gene regulatory activity (**Fig 2A**).

**Figure 2:**
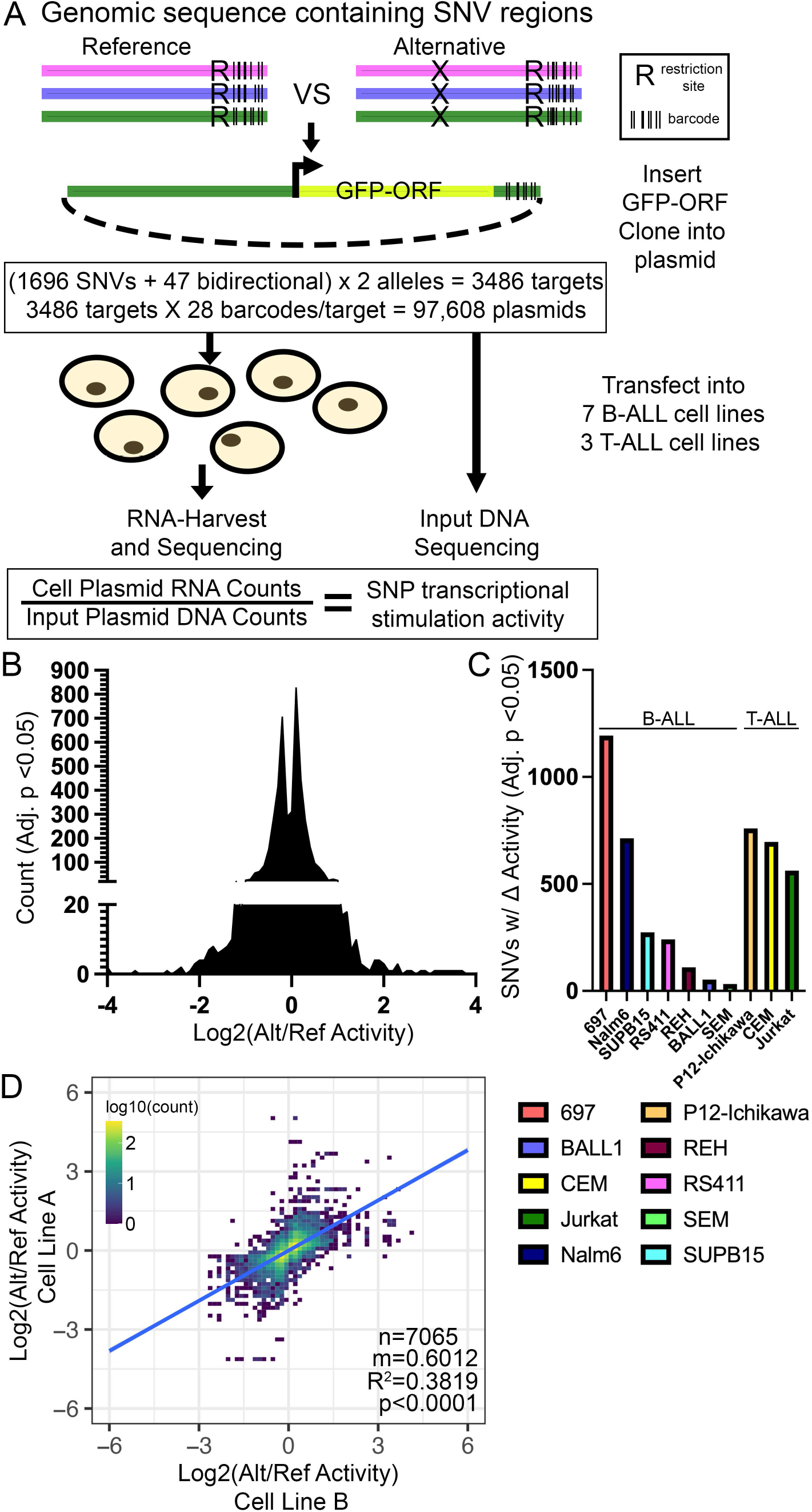
MPRA identifies regulatory variants with allele-specific effects on gene expression. **(A)** Diagram describing design of MPRA (also see **Methods**). **(B)** Distribution of significant changes in allele-specific transcriptional activity across all SNVs. **(C)** Number of MPRA SNVs showing significant (Adj. p<0.05) changes in allele-specific transcriptional activity in each ALL cell line. **(D)** Pair-wise linear correlation between changes in allele-specific transcriptional activity for all significant (Adj. p<0.05) changes across all cell lines. R^2^ correlation and p-value are provided.

Following transfection into 7 different B-cell precursor ALL (B-ALL; 697, BALL1, Nalm6, REH, RS411, SEM, SUPB15) and 3 T-cell ALL (T-ALL; CEM, Jurkat, P12-Ichikawa) human cell lines (n=4 transfections per cell line; 40 total), the transcriptional activity of each allele variant was measured by high-throughput sequencing to determine the barcode representation in reporter mRNA and compared to DNA counts obtained from high-throughput sequencing of the MPRA plasmid pool (**Fig 2A**). MPRA detected 4633 instances of significant differential activity between alleles across 91% (1538/1696) of regulatory variants across the 10 ALL cell lines tested (**Fig 2B-C, Sup File 2**). The 10 ALL cell lines showed substantial differences in the total number of regulatory variants harboring significant allele-specific activity (**Fig 2C**). Importantly, when comparing changes in allele-specific MPRA activity for each regulatory variant we found that significant changes in activity (adj. p<0.05) were highly correlated between ALL cell lines, with 87% concordance in allelic-specific activity, suggesting that significant MPRA hits were likely to be robust and reproducible between cell lines (**Fig 2D**). Allele-specific MPRA activities were also correlated using all pairwise cell line comparisons for each regulatory variant, irrespective of significance (**Sup Fig 1**). Importantly, 31 of the 35 positive control variants (i.e., ALL disease susceptibility-associated variants and variants in high LD) showed significant allelic effects in at least 1 cell line, and 10 showed significant and concordant allelic effects in at least 3 ALL cell lines, including two variants (rs3824662 at *GATA3* locus and rs75777619 at 8q24.21) directly associated with ALL susceptibility (*44, 49, 52*). The risk A allele at rs3824662 was associated with higher *GATA3* expression and chromatin accessibility and demonstrated significantly higher allele-specific activity in our MPRA (*44, 52*), thereby demonstrating that the MPRA could detect allelic effects identified by others. Overall, these data suggest that the chemotherapeutic drug sensitivity and patient treatment outcome SNVs tested were heavily enriched for functional regulatory variants with the potential to impact gene regulation.

### Identification of functional regulatory variants showing reproducible and concordant changes in allele-specific gene expression

To further focus on regulatory variants most likely to broadly impact gene regulation in ALL cells, we prioritized 556 variants with significant and concordant allele-specific activities in at least 3 ALL cell lines (i.e., functional regulatory variants; **Fig 3A-B, Sup File 3**). Most of these functional regulatory variants (318/556) mapped to accessible chromatin found only in primary ALL cell samples, underscoring the importance of incorporating chromatin architecture from primary ALL cells, and 54 functional regulatory variants mapped to transcription factor footprints in primary ALL cells (**Sup Fig 2**). However, because further functional investigation of variants in primary ALL cells is currently intractable, we focused on 210 functional regulatory variants that reside in open chromatin in an ALL cell line, and most of these variants (159/210; 76%) were also found in accessible chromatin in PDX and/or in primary ALL cells from patients (**Fig 3B-C**).

**Figure 3:**
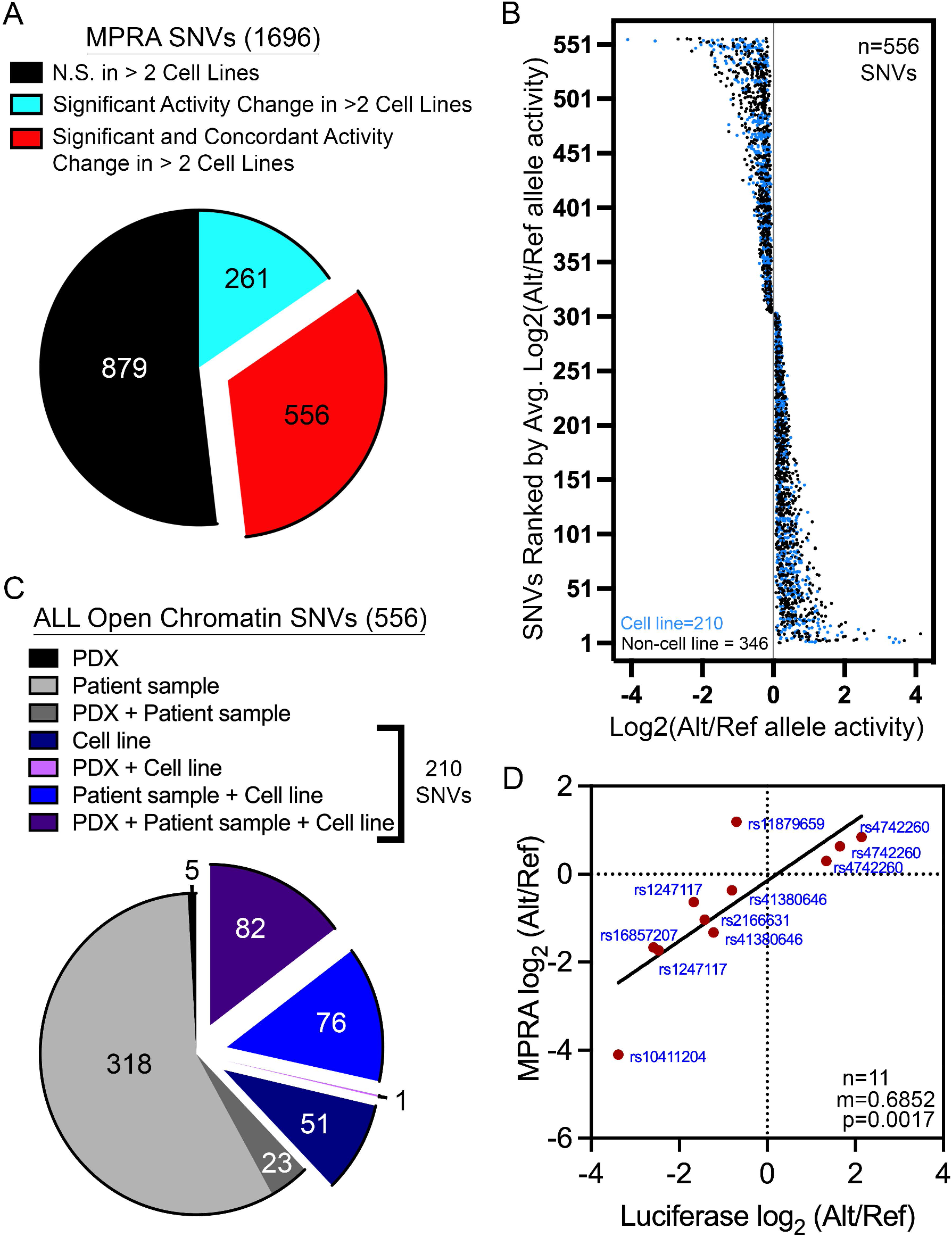
Identification of functional regulatory variants with reproducible and concordant effects in allele-specific stimulation of transcriptional activity. **(A)** 556 of the 1696 SNVs assayed are functional regulatory variants with reproducible (FDR<0.05 in >2 cell lines) and concordant (same directionality in >2 cell lines) changes in allele-specific activity. **(B)** Plot showing the distribution of log_2_-adjusted activity between alternative (Alt) and reference (Ref) alleles across 556 functional regulatory variants. **(C)** Pie chart shows how many functional regulatory variants map to open chromatin in diverse ALL cell models. 210 of the 556 functional regulatory variants are found in accessible chromatin sites that was identified in an ALL cell line. **(D)** Top hits from the 210 functional regulatory variants found in accessible chromatin in ALL cell lines were orthogonally validated by luciferase reporter assays. Data show significant correlation between the allele-specific effects detected by MPRA and dual-luciferase reporter assays.

For validation using traditional luciferase reporter assays, we prioritized these 210 functional regulatory variants based on allele-specific effect size and selected high-ranking SNVs with known eQTL status. Dual-luciferase reporter assays showed similar allele-specific changes in activity to that which was detected by MPRA, in validation of our MPRA analysis (**Fig 3D, Sup Fig 3**). In fact, a significant positive correlation (p=0.0017) was observed between the allelic effects detected by MPRA and luciferase reporter assays (**Fig 3D**). Together these analyses validated the robustness of our MPRA screen of functional regulatory variants and identified over 500 SNVs with reproducible and concordant allele-specific effects on gene expression.

### Association of functional regulatory variants with putative gene targets

We determined if the 210 functional regulatory variants found in accessible chromatin sites in ALL cell lines were directly associated with target gene regulation. While 34 functional regulatory variants were localized close (+/-2.5kb) to nearby promoters (**Fig 4A, Sup File 4**), 176 variants were promoter-distal, and therefore likely to map to CREs with unclear gene targets (**Fig 4A**). We therefore performed promoter CHiC in 8 of 10 ALL cell lines used in MPRA and determined that 19 of the 176 functional regulatory variants showed evidence of connectivity to distal promoters in the same cell line where allele-specific MPRA activity and chromatin accessibility were detected (**Fig 4A, Sup File 4**). In prioritizing functional regulatory variants, we were interested in the gene regulatory impacts of variants at TSS-proximal promoter-associated versus TSS-distal promoter-connected CREs. Interestingly, we found that SNVs found in open chromatin at TSS-distal sites with promoter connectivity showed higher allele-specific changes in MPRA activity than those at promoters (**Fig 4B**). Amongst the TSS-distal promoter-connected functional regulatory variants, we found that distal intergenic and intronic SNVs showed significantly higher allele-specific activity than those in UTRs (**Fig 4C**). These data suggest that the most robust allelic effects attributable to these regulatory variants are likely to occur at distal intergenic and intronic sites >2.5kb from the TSS of the target gene.

**Figure 4:**
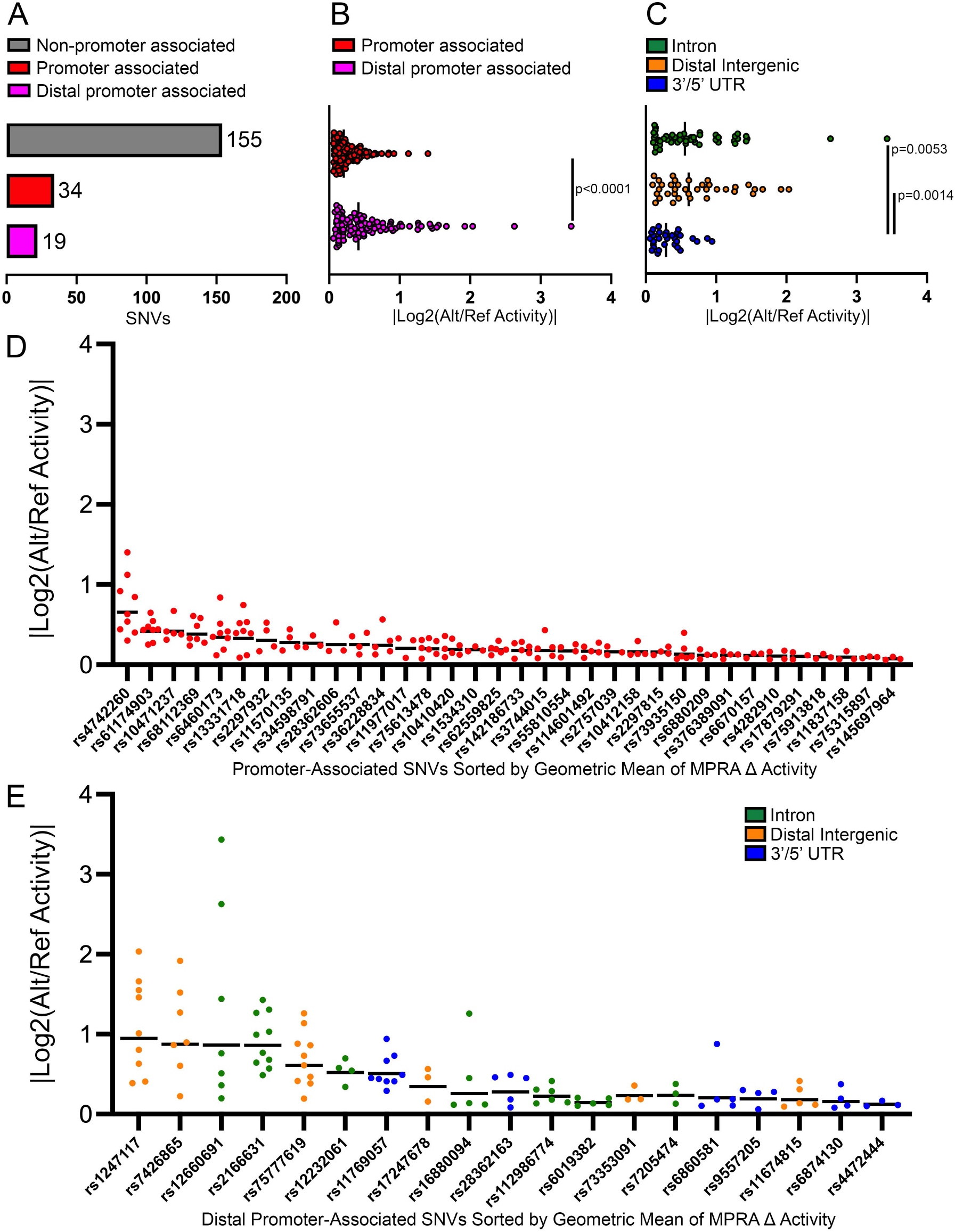
Promoter CHiC identifies target genes of functional regulatory variants. **(A)** Data show the number of functional regulatory variants mapping to open chromatin in cell lines that associate directly with promoters or that are distally promoter-connected via promoter CHiC. **(B)** MPRA data show promoter-connected functional regulatory variants in accessible chromatin exhibit stronger effects on allele specific activity than promoter-associated functional regulatory variants. **(C)** Amongst distally promoter-connected functional regulatory, variants that map to intronic and distal intergenic sequences showed greater activity than those in UTRs. **(D-E)** Data show the ranked allele-specific activity distribution of MPRA data for **(D)** promoter-associated functional regulatory variants and **(E)** distally promoter-connected functional regulatory variants.

Next, we ranked TSS-proximal promoter-associated and TSS-distal promoter-connected functional regulatory variants by the geometric mean of their significant MPRA data to account for the magnitude of allele-specific activity and the reproducibility of a significant change across ALL cell lines (**Fig 4D-E**). This analysis identified rs1247117 as the most robust functional regulatory variant to pursue for mechanistic understanding (**Fig 4E**).

### rs1247117 determines PU.1 binding and impacts sensitivity to vincristine

We pursued functional validation of rs1247117 based on its highest ranking by geometric mean of MPRA allelic effect. rs1247117 is in high LD with two GWAS sentinel variants (rs1312895, r^2^=0.99; rs1247118, r^2^=1) that are associated with persistence of MRD after induction chemotherapy (*3*). This functional regulatory variant maps to a distal intergenic region near the *CACUL1* gene, for which it is an eQTL for in EBV-transformed lymphocytes (*37*). However, rs1247117 also loops to the *EIF3A* promoter in Nalm6 B-ALL cells (**Fig 5A**). We therefore explored how this accessible chromatin site might recruit transcriptional regulators that would depend on the allele present at rs1247117. For this, we first performed ChIP-seq for RNA pol II and H3K27Ac which further confirmed that rs1247117 is associated with an active CRE in Nalm6 cells (**Fig 5A**). Through an examination of the underlying DNA sequence spanning rs1247117, we found that the reference guanine (G) risk allele at rs1247117 resides in a PU.1 transcription factor binding motif that is disrupted by the alternative adenine (A) allele. Although the risk G allele is the reference allele, the alternative A allele is more common in human populations. Supporting PU.1 binding at this location, accessible chromatin profiling in primary ALL cells identified an accessible chromatin site and PU.1 footprint spanning rs1247117 in diverse ALL samples (**Sup Fig 4A-B**). Significantly greater chromatin accessibility at rs1247117 was also observed in heterozygous (GA) patient samples compared to patient samples homozygous for the alternative A allele (**Sup Fig 4C**), and the G allele at rs1247117 harbored significantly greater ATAC-seq read count compared to the A allele (**Sup Fig 4D**). Importantly, we determined that PU.1 was bound at this site in Nalm6 cells using CUT and RUN (*57*) (**Fig 5A-B**).

**Figure 5:**
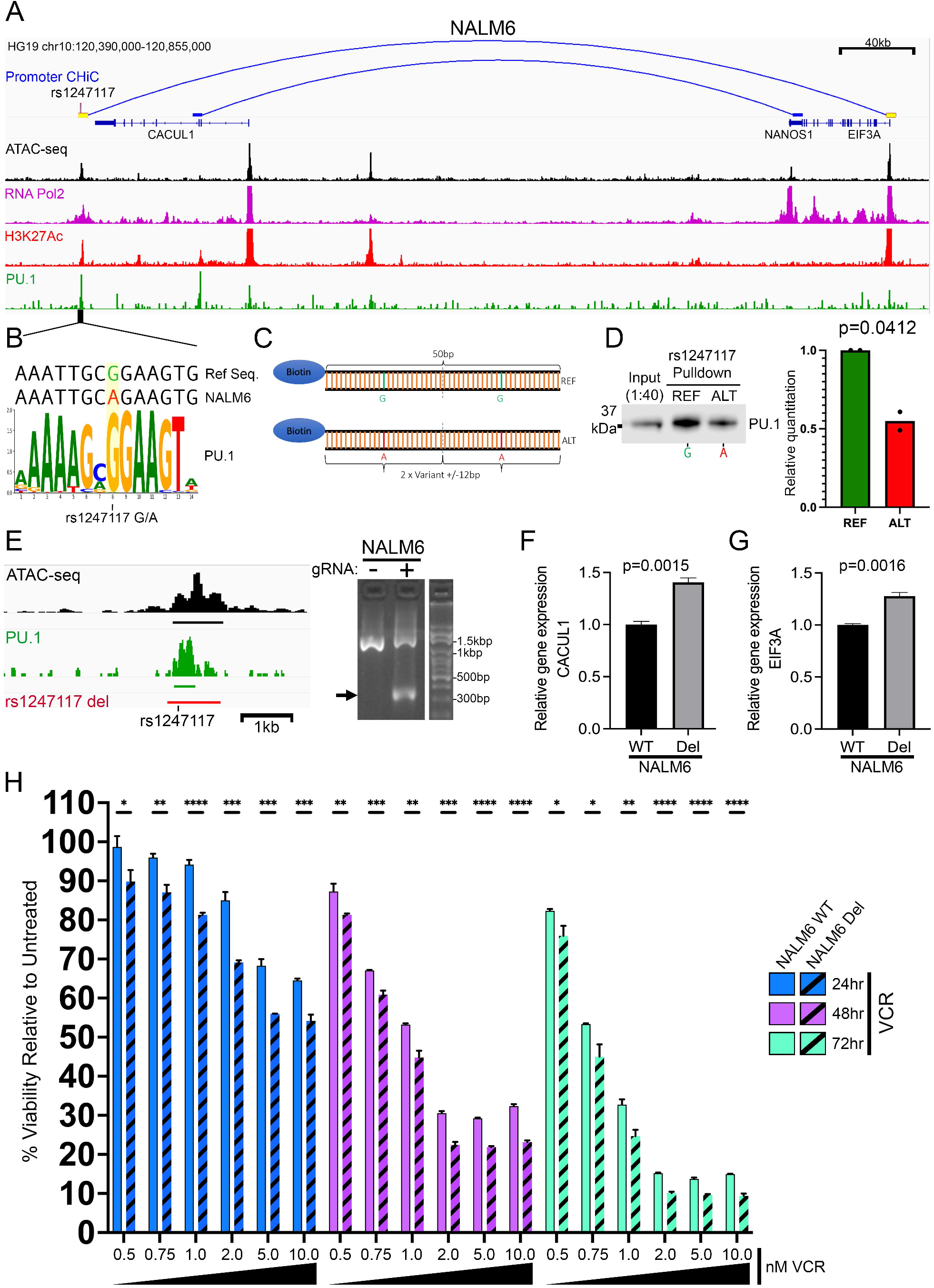
Functional exploration of rs1247117 in B-ALL cells. **(A)** IGV genome browser image in Nalm6 cells showing the genomic context, chromatin accessibility, and *EIF3A* promoter connectivity using promoter CHiC of the top functional regulatory variant, rs1247117, with the highest allele-specific MPRA activity. Genomic binding profiles are also shown for RNA polymerase II (RNA Pol2), histone H3 lysine 27 acetylation (H3K27Ac) and PU.1. **(B)** rs1247117 lies in a PU.1 binding motif. The human genome reference sequence, Nalm6 genome sequence, location of rs1247117 and PU.1 position weight matrix is shown. **(C)** Design of biotinylated DNA-probes for *in vitro* rs1247117 pulldown. **(D)** Biotinylated DNA pulldown shows rs1247117 allele-dependent enrichment of PU.1 binding. Blot shown is representative of two independent experiments. P-value from densitometric quantification of two blots is shown. **(E)** Diagram on the left showing the genomic context of the rs1247117 CRE deletion in Nalm6 cells in relation to chromatin accessibility, PU.1 binding and rs1247117. Black bar represents ATAC-seq peak, green par represents PU.1 peak, and red bar represents region deleted using CRISPR/Cas9 genome editing. Gel shows validation of deletion using primers flanking deleted region. Arrow points to PCR fragment with deletion in heterogeneous Nalm6 cell pools harboring deletion compared to wild-type parental Nalm6 cells. **(F-G)** *CACUL1* **(F)** and *EIF3A* **(G)** expression is upregulated upon deletion of the CRE containing rs1247117. RT-qPCR data show the mean +/- SEM of three independent experiments. **(H)** Drug sensitivity data comparing survival of wild-type parental Nalm6 cells and Nalm6 cells with rs1247117 CRE deletion after vincristine (VCR) treatment for 24 (n=3), 48 (n=3) and 72 (n=3) hours. Vincristine concentration is provided below. *, p<0.05; **, p<0.01; ***, p<0.001; ****, p<0.0001. Data show the mean +/- SEM relative to untreated cells.

Nalm6 cells contain the alternative A allele that disrupts the PU.1 motif at rs1247117, yet our data suggests that this site still binds PU.1 (**Fig 5A-B**). This led us to hypothesize that PU.1 binding affinity for the PU.1 motif surrounding rs1247117 would be strengthened by the risk G allele. Therefore, we designed biotinylated DNA probes containing two tandem 25-bp regions centered on reference G or alternative A allele-containing rs1247117 to test this hypothesis (**Fig 5C**). Using biotinylated probes we performed an *in vitro* DNA-affinity pulldown from Nalm6 nuclear lysate and found that while PU.1 was indeed bound to the alternative A allele, PU.1 was more robustly bound to the reference G allele at rs1247117 (**Fig 5D**). These data suggest that the risk G allele increases the affinity of PU.1 binding at rs1247117 relative to the alternative A allele.

We were next interested in how allele-specific PU.1 binding was related to the expression of nearby putative target genes. Because rs1247117 is a known eQTL associated with *CACUL1* expression in EBV-transformed lymphocytes and our promoter CHiC data demonstrated connectivity between rs1247117 and the promoter of *EIF3A*, we asked if the expression of these two genes was altered by deletion of the CRE containing rs1247117 in Nalm6 cells. Using CRISPR/Cas9 genome editing, we made a heterogeneous pool of Nalm6 cells harboring a deletion of the CRE containing rs1247117 (**Fig 5E**). We found that CRE deletion resulted in a significant up-regulation of both *CACUL1* and *EIF3A* (**Figure 5F-G**), suggesting an inverse relationship between PU.1 binding and transcription of associated genes at this locus. Importantly, this observation is concordant with GTEx eQTL data showing that the risk G allele harboring stronger PU.1 binding is associated with down-regulation of *CACUL1* in EBV-transformed lymphocytes (*37*).

Because the risk G allele at rs1247117 was also associated with vincristine resistance in primary ALL cells from patients (*3*), we additionally sought to determine the impact of the CRE deletion containing rs1247117 on cellular response to vincristine treatment. We hypothesized that because the risk G allele is associated with enhanced PU.1 binding and resistance to vincristine, complete disruption of PU.1 binding in Nalm6 cells harboring the CRE deletion would show increased sensitivity to vincristine relative to parental Nalm6 cells. As predicted, Nalm6 cells with the CRE deletion exhibited significantly increased sensitivity to vincristine across a range of concentrations after 24, 48, and 72 hours of treatment (**Fig 5H**). Collectively, these data suggest that a functional regulatory variant alters the binding affinity of a key transcription factor, PU.1, and disruption of PU.1 binding at this locus impacts vincristine sensitivity in ALL cells.

## DISCUSSION

Using MPRA, we systematically interrogated the functional effects of inherited noncoding variation associated with relapse, persistence of MRD after induction chemotherapy and/or *ex vivo* chemotherapeutic drug resistance in childhood ALL. We refined our search to regulatory variants that were found in accessible chromatin sites in 158 ALL cell models, including primary ALL cells from patients, PDXs and ALL cell lines, as those noncoding regions were likely to be participating in transcriptional regulation. Using MPRA we identified 556 functional regulatory variants showing reproducible and concordant changes in an allele-specific gene regulatory activity. To further explore the impact of these variants on gene regulation in ALL cell lines, we selected a subset of functional regulatory variants from MPRA that were within an accessible chromatin site in an ALL cell line. We overcame difficulties in associating promoter-distal functional regulatory variants with gene targets using promoter CHiC, and found 19 variants with robust looping to a distal promoter, as well as 34 promoter-associated functional regulatory variants.

We identified rs1247117 as the top functional regulatory variant showing the highest geometric mean of differential transcription activity, which was identified in 9 of 10 ALL cell lines assayed by MPRA. We found that the allele present at rs1247117 was determinant of PU.1 transcription factor binding, with the risk G allele leading to greater chromatin accessibility and PU.1 binding affinity. Interestingly, the allele-specific activities as measured by MPRA and traditional dual-luciferase reporter assays suggest that the reference G allele at rs1247117 stimulates transcription more than the alternative A allele, and we suspect this is driven by greater PU.1 binding affinity in these episomal assays. However, our endogenous genetic manipulation that disrupted PU.1 binding altogether at this locus in Nalm6 cells suggest that *CACUL1* and *EIF3A* expression are driven inversely to PU.1 binding. Corroborating these endogenous findings, GTEx eQTL data suggests that the risk G allele harboring great PU.1 affinity is associated with reduced *CACUL1* expression. Moreover, complete disruption of PU.1 binding resulted in greater sensitivity to vincristine, which is consistent with the risk G allele contributing to both greater PU.1 binding affinity and vincristine resistance in primary ALL cells from patients. This discrepancy may be due to the ability of PU.1 to act in an activating or repressing manner on gene expression dependent on its genomic context and other transcriptional regulators present (*58, 59*). While not addressed within the scope of this work, our hypothesis is that MPRA and luciferase reporter assays, which are episomal and utilize a non-native minimal promoter, detected transcription activating PU.1 activities rather than the PU.1 repressive activities we detect within the endogenous locus. Collectively, these observations stress the importance of performing subsequent functional follow-up experimentation within an endogenous sequence context.

Although the risk G allele at rs1247117 is associated with decreased *CACUL1* expression, increased risk of MRD after induction chemotherapy and vincristine resistance, it remains unclear why *CACUL1* expression might impact vincristine efficacy. *CACUL1* expression has been correlated with cell cycle progression, and others have shown that *CACUL1* knockdown leads to cell cycle arrest at the G1/S checkpoint (*60*). However, vincristine can act on microtubules to rapidly kill cells during G1 and later the mitotic spindle to arrest cells during metaphase, so increased *CACUL1* expression may facilitate cell cycle progression, thus increasing the rate at which metaphase mitotic spindles are disrupted by vincristine (*61*).

Our studies also identified *EIF3A* as a target gene for rs1247117 through long-distance promoter looping. *EIF3A* expression has been previously linked to chemotherapeutic sensitivity in both melanoma and lung cancer (*62, 63*). EIF3A expression led to decreased phosphorylation of ERK, supporting the effect of vemurafenib-induced MAP kinase signaling blockade, while EIF3A loss led to sustained activation of ERK and therapeutic resistance (*62*). Interestingly, ERK activation is important in G1/S progression, and therefore it follows that EIF3A-dependent inhibition of ERK may support the rapid killing of cells in G1 phase shortly after initial vincristine treatment (*61, 64*). This notion is supported by significantly greater sensitivity to vincristine of ALL cells harboring greater *EIF3A* expression through disruption of a distal CRE after just 24 hours of treatment.

The regulatory variants assayed in this study were originally discovered from GWAS in patient samples, and most of our functional regulatory variant hits from MPRA were present in accessible chromatin sites found only in primary ALL cells from patients. Consequently, these data highlight both substantial differences in the chromatin landscape between immortalized cell lines and primary cells and a limitation of our study that relied on the functional exploration of the top regulatory variant in an ALL cell line model. An optimal approach would be to validate top functional regulatory variants in patient samples; however, this is not currently feasible due to the limited duration of patient sample viability in culture for genetic manipulation. Nonetheless, future implementation of promoter CHiC in patient samples can be used to map gene connectivity of promoter-distal functional regulatory variants found only in primary cells, and these gene targets can then be genetically disrupted in ALL cell line models for functional validation.

This translational work represents the largest functional investigation of inherited noncoding variation that is associated with diverse pharmacological traits in ALL. Our study identified hundreds of functional regulatory variants with significant, reproducible, and concordant allele-specific effects on gene expression, and further connected gene regulatory disruptions to differences in chemotherapy response through alterations in antileukemic drug sensitivity in ALL cells. Collectively, these data support the importance of noncoding, gene regulatory disruptions in the pharmacogenomics of ALL treatment. The further functional investigation of these regulatory variants and the discovery of additional inherited variants impacting therapeutic outcome can be used by clinicians to tailor therapies based on a patient’s unique genetic makeup through precision or personalized medicine.

## MATERIALS AND METHODS

### Patient samples and consent

All patients or their legal guardians provided written informed consent. The use of these samples was approved by the institutional review board at St. Jude Children’s Research Hospital. Patient samples were obtained from: St. Jude Children’s Research Hospital (Memphis, Tennessee) Total Therapy XVI protocol (TOTXVI, NCT00549848) and Total Therapy XVII protocol (TOT17, NCT03117751); Eastern Cooperative Oncology Group (ECOG), The Alliance for Clinical Trials in Oncology, MD Anderson Cancer Center (Houston, Texas) or the University of Chicago (Chicago, Illinois).

### *Ex vivo* drug resistance assays in primary ALL cells

Primary leukemia cells were isolated from the bone marrow or peripheral blood of newly diagnosed ALL patients from St. Jude Total Therapy XVI protocol (TOTXVI, NCT00549848) and tested for antileukemic drug sensitivity by a 96-hour MTT assay using a range of drug concentrations, as previously described (*20, 21*). Primary ALL cells were treated with prednisolone (n=320), dexamethasone (n=312), bacterially derived L-asparaginase (n=335), vincristine (n=323), 6-mercaptopurine (n=344) and 6-thioguanine (n=325). Following drug treatment, the lethal concentration resulting in 50% viability (LC_50_) was calculated for each patient sample.

### Nalm6 vincristine sensitivity assays

Drug viability assays were performed as previously with slight modification (*22*). Nalm6 parental cells (WT) and rs1247117 CRE deletion Nalm6 cells (Del) were seeded at 20,000 cells per well in a 96-well plate and co-treated with the indicated concentrations of vincristine (Hospira, 61703-0309-16). Following the indicated duration of incubation, cell viability was measured using the CellTiter-Glo® 2.0 Cell Viability Assay (Promega, G9243). The luminescence was measured using a BioTek Cytation1 cell imaging multimode reader (Agilent). The obtained values were normalized and plotted as % of control. All the experiments were performed in 3 biological replicates having 4-6 technical replicates in each group. Data was plotted as Mean +/- SD.

### Selection of SNVs impacting treatment outcome in patients from published GWAS

We chose 13 relapse SNVs with p<1×10^−5^ from (*4*), 19 ancestry-specific SNVs relapse SNPs associated with relapse in both discovery and replication ALL patient cohorts (p<0.05) from (*5*) and 3 SNVs associated with persistence of MRD with p<1×10^−6^ from (*3*) (n=35). In addition, we chose all SNVs (n=126) from these GWAS with nominal genome-wide association (p<0.05) but that were additionally associated with *ex vivo* drug resistance phenotypes in primary ALL cells from patients (p<0.05) enrolled in St. Jude Total Therapy XIIIB, XV and/or XVI protocols (TOTXIIIB, TOTXV and TOTXVI).

### Genotyping in primary ALL cells

DNA was extracted from the ALL cells of bone marrow or peripheral blood samples from patients using the Blood and Cell Culture DNA kit (Qiagen). Genotyping was performed using the Affymetrix GeneChip Human Mapping 500K set or the SNP 6.0 array (Affymetrix). Genotypes were called BRLMM algorithm in the Affymetrix GTYPE software (http://www.affymetrix.com/products/software/specific/gtype.affx) as previously described (*65*). We excluded SNVs for call rates <95% among patients or minor allele frequencies <1%.

### Gene expression profiling in primary ALL cells

Total RNA from primary ALL cells was isolated using RNAeasy Mini kit (Qiagen) and mRNA sequencing using an Illumina HiSeq platform was performed by the Hartwell Center for Bioinformatics and Biotechnology at St. Jude Children’s Research Hospital.

### Quantitative real time PCR (qPCR)

Nalm6 parental (WT) and rs1247117 deleted Nalm6 cells were cultured in RPMI 1640 media. 10 million cells were collected from each group in triplicates and resuspended in RLT/BME mixture for total RNA extraction. RNA was isolated using RNeasy Mini Kit (Qiagen #74104). Complimentary DNA synthesis was done using the High-Capacity RNA-to-cDNA Kit (Applied Biosystems #4387406). RT-PCR reactions were prepared using TaqMan Fast Advanced Master Mix (Applied Biosystems #4444557) and TaqMan Gene Expression Assays (Thermo) (CACUL1: Hs00403870_m1, EIF3A: Hs01025769_m1, TBP (endogenous control, Hs00427620_m1). The samples were run on a QuantStudio 3 Real-Time PCR Instrument using the recommended TaqMan Fast Advanced Master Mix PCR conditions.

### Dual-luciferase reporter assays

A 300-bp of sequence centered on reference or the alternative allele of rs1247117, rs10411204, rs4742260, rs12660691, rs2166631, rs11879659, rs41380646, rs16857207 was cloned upstream of the minimal promoter into the pGL4.23-basic vector. Sequences used in luciferase reporter experiments are shown in **Supplemental File 5**. Nalm6, SUPB15, REH, and 697 cells (10 million cells per replicate, 60 μg plasmid DNA and 6 μg pRL-TK control vector) were used for transfection. Using Neon Transfection system (Thermo Fisher Scientific, MPK5000), the constructs were co-transfected with renilla plasmid to enable normalization of the luciferase signal. 24 h post-transfection, firefly luciferase and renilla luciferase activity was measured using Dual Luciferase Reporter Assay System (Promega, E1960) on a BioTek Cytation1 cell imaging multimode reader (Agilent). The ratio of firefly luciferase to renilla luciferase activity readings reflect the luciferase activity of the reference allele relative to the alternative allele. All experiments were performed in 10 samples from each replicate and repeated 2-3 times.

### Chromatin accessibility mapping in ALL cell models

Fast-ATAC in fresh primary ALL cells from patients (n=120), PDXs (n=3) and in a subset of ALL cell lines (BALL1, CEM, Jurkat and P12-Ichikawa) was performed on 10,000 cells as described in (*22, 66*). Paired-end Illumina next-generation sequencing of Fast-ATAC libraries was performed at the Hartwell Center for Bioinformatics and Biotechnology at St. Jude Children’s Research Hospital. Data were analyzed as in (*22*). For cryo-preserved primary ALL cells from patients and for B-ALL cell lines, Fast-ATAC data was obtained from the Gene Expression Omnibus (GSE161501 and GSE129066).

### Massively parallel reporter assays

#### MPRA Oligo design

Oligo libraries were designed by following previous work with modified protocols (*27, 31, 67, 68*). MPRA oligos ordered from Agilent (230 bp) were structured as follows: 5’-Primer1-enh-Kpnl-Xbal-barcode-primer2-3’ where primer1 and primer2 are universal primer sites, *enh* denotes the 175bp variant containing region to test for enhancer activity, Kpnl and Xbal denote recognition sequences for cut sites, and *barcode* denotes 10-bp tag sequence (see **Sup File 6**). Agilent oligos were resuspended in 100 ul nuclease free water. All 10-bp barcodes for each variant allele used in MPRA are provided in **Supplemental File 7**.

#### MPRA Plasmid Cloning-Input (DNA) Library construction

For plasmid cloning, oligo libraries were amplified by 20 cycles of emulsion PCR (Micellula DNA Emulsion & Purification Kit #E3600, EURx Molecular Biology Products) using Herculase II fusion DNA polymerase (#600675, Agilent), forward and reverse primers (see Sup File 3) to introduce SfiI restriction enzyme sites (GGCCNNNNNGGCC) (NEB) and homology arms to the pMPRA1 plasmid (Cat: #49349, Addgene). Purified PCR products were separated on a 2-4% agarose gel to verify the expected amplification size of 281bp. The pMPRA1 backbone vector was Sfil digested overnight and size selected on a 1% agarose gel. Vector backbone gel extraction was done with the Qiagen gel extraction kit and QIAquick PCR purification kit (28706×4 and 28104). Gibson assembly was used to clone oligos into vector using 79 ng of inserts, 100 ng of digested vector, and 20 ul Gibson assembly 2x master mix (# E2611S, NEB). The reaction was purified using MinElute PCR purification column (Qiagen), and drop analysis was performed with Millipore filters (Type VSWP 0.25 um Millipore #VSWP02500). For the transformation step, we aimed to obtain 10x CFU bacterial cells than the distinct promoter-tag combinations (unique sequences) in the oligo library. We transformed the Gibson assembly reactions into MegaX DH10B electrocompetent bacteria (#C6400-03, Invitrogen) using GenePulser II electroporator (Bio-Rad). Plasmids were extracted with Qiagen Maxi Prep Kit. For quality control studies, an aliquot of the isolated plasmid library was digested with SfiI and run on 1 % agarose gel to confirm the presence of inserts. To generate linear enhancer-barcode backbone sequences for reporter insertion, 2 ug of plasmid was digested with KpnI/XbaI. A minimal promoter + truncated eGFP was then ligated to the linearized enhancer-barcode backbone and purified using the Qiagen MilEute PCR purification kit. The ligation was then transformed into 1 vial of MegaX electrocompetent bacteria (#C6400-03, Invitrogen) as before. Plasmids were then extracted using a Qiagen Maxiprep kit as before and the elution was verified as a single size band by gel electrophoresis.

#### MPRA library transfection and sequencing

MPRA plasmid library (10 μg) transfections were done in 10 ALL cell lines having at >95% cell viability (45 million cells x 4 replicates x 10 cell lines) using electroporation with the Neon Transfection system (Thermofisher). Next day, RNA-was harvested using the RNeasy plus mini kit (Cat: #74134, Qiagen) using 4 columns per sample. Once the RNA was isolated, we performed additional DNase digestion using RQ1 RNase-free DNase (Cat: # M6101, Promega). All tubes from the same replicates were combined and added 1 volume of 70% ethanol to the combined lysate and mixed well by pipetting. The DNase treated RNA was again purified with RNeasy mini kit and eluted in 60 ul RNase free water. Total RNA was quantified using DeNovix Ds-11 FX instrument. We yielded 16μg-112μg of total RNA from each replicate depending on the cell line used. mRNA purification was performed using Dynabeads mRNA purification kit (Cat: #61006, Invitrogen). mRNA concentration was measured using Qubit HS RNA (Cat: #Q32852, Invitrogen). We yielded from 0.75-2 ug of mRNA in average from each replicate. cDNA was synthesized using three primers (2 uM) cDNA P1, cDNA P4, and cDNA 6, with the Superscript III first-strand synthesis system (Cat: #18080051, Invitrogen) (see **Sup File 6**).

Final multiplexing of 50ng cDNA and input plasmid DNA (4 aliquots of MPRA plasmid pool that were independently prepared for next-generation sequencing) was carried out using Q5 Hot Start 2x Master Mix (NEB #M0494S), index primers, and Multiplexing primer 1 for 15 cycles of PCR. The reactions were size selected using AMPure XP beads (Cat: #A63881, Beckman Coulter, Indianapolis, IN) and eluted in 20 ul nuclease free water. The final library concentration was measured using Qubit DNA HS. 20-40 ng of each library was sequenced on the Illumina NovaSeq (200 million x 150bp paired-end reads per sample) at the Hartwell Center for Bioinformatics and Biotechnology at St. Jude Children’s Research Hospital.

#### MPRA sequencing analysis

Following next-generation sequencing, the MPRA sequence data was trimmed to contain only barcode sequences without allowing for any mismatches and read counts were determined for all barcodes. To identify significant allele-specific effects mpralm (*69*) was performed on RNA and DNA barcode counts after merging RNA or DNA counts across all barcodes for each allele.

#### Promoter capture Hi-C

Arima promoter capture HiC (Arima: A510008, A303010, A302010) was performed according to the manufacturers provided instructions using unspecified proprietary buffers, solutions, enzymes, and reagents. Briefly, 10 million ALL cells were harvested, suspended in 5ml RT PBS which was brought to 2% formaldehyde by adding 37% methanol-stabilized paraformaldehyde for a 10-minute fixation. The amount of fixed cell suspension equal to 5μg of cell DNA was used for HiC. Cells were lysed with Lysis Buffer and conditioned with Conditioning Solution before their DNA was digested in a cocktail consisting of Buffer A, Enzyme 1, and Enzyme 2. The digested, fixed chromatin was biotinylated using Buffer B and Enzyme B before being ligated using Buffer C and Enzyme C. The fixed, biotinylated, ligated DNA was then subjected to reversal of crosslinking and digestion of proteins before being purified. 100ul containing 1500ug of purified large proximally ligated DNA was fragmented for 24 cycles (30s on/ 30s off) using a Diagenode Bioruptor Plus bath sonicator. The fragmented DNA was then subjected to two-sided size selection targeting fragments between 200-600bp using AMPure XP DNA purification beads. Size selected DNA was then subjected to biotin enrichment using T1 streptavidin beads. Bead bound, enriched HiC DNA was then subjected to Arima library prep. Briefly, the sample underwent end repair followed by adapter ligation, at which point the sample was then subjected to 10 cycles of PCR amplification. The library DNA was then purified using AMPure XP DNA purification beads. The HiC library was then subjected to Arima promoter capture enrichment. The library was precleared of biotinylated DNA using T1 streptavidin beads before being subjected to promoter enrichment with biotinylated RNA probes. After washing, the captured fragments were then amplified an additional 13 PCR cycles. These libraries were submitted for deep sequencing on an Illumina Nova-seq where >200M 150bp paired-end reads were obtained. Analysis of Promoter capture HiC data was performed using the Arima CHiC pipeline (v1.5, https://github.com/ArimaGenomics/CHiC). Briefly, this pipeline uses HiCUP v0.8.0 for mapping and quality assessment of promoter capture HiC data and CHiCAGO to identify significant looping interactions in the promoter capture HiC data using 5kb resolution and adj. p < 0.05 (*70, 71*).

#### PU.1 *in vitro* binding affinity assay

DNA pulldown assay was adopted from previous article and performed using a modified protocol (*72*). Briefly, biotinylated ssDNA probes were ordered via custom synthesis from IDT with their non-biotinylated reverse complement sequences. The DNA probe sequences used in the experiment are listed in **Supplemental File 8**. The probes containing reference and alternative alleles featuring the nucleotide of interest (rs1247117) and its flanking +/-12 bp nucleotides were arranged side-by-side in tandem for a total of 50 bp each. Probes were annealed by combining biotinylated probes and the non-biotinylated reverse complement at 1.5M excess (50μM:75μM) with an equal volume of 2x annealing buffer (10 mM Tris pH 8.0, 100 mM NaCl, 2 mM EDTA) and incubating at 98C for 10 minutes before cooling at RT overnight. To isolate nuclear lysate, we washed 75 million cells in 5 mL PBS and pelleted at 500xg for 3 min at RT. The cells were resuspended in 2 mL of homogenization buffer (1M KCl, 1M MgCl2, 1M HEPES, 0.5 M EGTA, 1x Halt protease inhibitors, Thermo Fisher 78429) and passed through 26 G needle 10x. The nuclei were pelleted, and the supernatant was discarded. The nuclei were gently washed with an additional 1ml of homogenization buffer, pelleted, and the supernatant discarded again. Washed nuclei were suspended in 300 μL of SKT buffer (1M HEPES, 1M MgCl2, Glycerol, 1 M KCl, EDTA, 0.1% triton). To extract nuclear proteins 33ul of 3M NaCl was added and samples were vortexed every 2-3 min on ice for the next 20 min. Insoluble nuclear material was pelleted at 13000 rpm for 10 min at 4°C. To make a nuclear lysate master mix 1100μg of nuclear lysate was transferred to a fresh tube with 22μg nonspecific DNA (11μg Poly (dI-dC), (Thermo Fisher 20148E) + 11μg Poly(dA:dT), (Cell Signaling Technologies)) and brought to a final volume of 1320μL with protein binding buffer (PBB, 150 mM NaCl, 0.25% NP40, 50 mM Tris pH 8.0, 1 mM DTT and EDTA free protease inhibitors (Roche). Reference and Alternative allele annealed biotinylated DNA probes (500pMol) were prebound to 20ul Streptavidin T1 Dynabeads (#65601, Thermo) in 600μL DNA binding buffer (DBB, 1 M NaCl, 0.05% NP40, 10 mM TRIS, pH 8.0 and 1 mM ETDA) rotating at 4°C for 30 min. Streptavidin T1 beads bound to probes were washed 1x DBB, 2x PBB on ice and 600 uL of nuclear lysate master mix (prepared above) was added to reference or alternative allele bead-bound probe tubes. Lysate and probe-bound beads were rotated for 90 min at 4°C and washed 3x PBB, and 2x PBS by pipetting up and down 5x each wash. Proteins were eluted in 40 ul 1x LDS sample buffer (+10% BME) (Cat: #NP0007, Invitrogen) by heating at 99.9°C in a thermal mixer at 1200 rpm for 10 min (#13687712, Thermo). Samples were subsequently assessed by western blot as previously described (*22*) using anti-rabbit PU.1 antibody (#2258S, Cell Signaling Technology).

#### ChIP-seq

RNA polymerase II ChIP-seq data were generated by first fixing 20 million Nalm6 cells in 1% formaldehyde (diluted from sigma F87750) at room temp for 10 minutes. Crosslinking was stopped with the addition of 2.5M glycine to a concentration of 0.125M, and the cells were then washed in ice-cold PBS. 5μg anti-RNA polymerase II CTD repeat YSPTSPS (phospho S5) antibody [4H8] (ab5408, lot: GR3264797-1) was prebound to 200ul of protein G dynabeads (Invitrogen 10003D) overnight in 0.5% BSA in PBS. Fixed cell pellets (20M cells) were suspended in 1ml Farnham lysis buffer (5mM PIPES pH 8, 85mM KCl, 0.5% NP40, 1x protease inhibitors (Roche 11836170001)) and passed through a 18G needle 10x. Nuclei were pelleted and resuspended in 275ul of RIPA buffer (1x PBS, 1% NP40, 0.5% Sodium Deoxycholate, 0.1% SDS, 1x protease inhibitors) and sonicated on high power in 1.5ml tubes for 25 minutes (30s on/ 30s off) using a Diagenode Bioruptor Plus. 5% Input samples were taken from sonicated material and the remaining sonicated material was added to the pre-bound antibody/protein G beads to rotate overnight at 4C. The next day the supernatant was discarded, and the beads were washed 5x with ice cold LiCl buffer (100mM Tris pH 7.5, 500mM LiCL, 1% NP40, 1% sodium deoxycholate) and 1x with ice cold TE buffer (10mM Tris pH 7.5, 1mM EDTA). Samples were eluted from the washed beads using room temperature IP elution buffer (1% SDS, 0.1 M NaHCO_3_) at 65C for 1hr, vortexing every 15 minutes. The elution was then incubated at 65C overnight to reverse crosslinks. The next day DNA was purified using the QIAquick PCR purification kit (Qiagen 28104). DNA quantification was performed using the PicoGreen assay (Molecular Probes, Eugene, OR, P-7581). Sequencing libraries were generated from ChIP and input DNA by using the KAPA Hyper Prep kit (Roche, Basel, Switzerland, # 7962363001) according to the included manufacturer’s specifications, and quality was determined by using the Agilent TapeStation with D1000 screentape. Then, >50M 50-bp paired-end reads per sample were generated on the NovaSeq 6000. Reads were quality checked using fastqc (v0.11.5) and trimmed using trimgalore (v0.4.4) before being mapped to the hg19 reference genome using bowtie2 (v2.2.9). Sam files were converted to bam format using samtools (v1.2), which were sorted using picard (v1.141). Duplicates were removed using picard and mitochondrial reads were removed using samtools. For visualization, bam files from replicates were merged using samtools and converted to bigwig format using deeptools (v3.5.0). For peak calling, we used macs2 (v2.1.1), and only considered peaks called in both samples. H3K27ac ChIP-seq data in Nalm6 cells was obtained from the Gene Expression Omnibus (GSE161501).

#### PU.1 CUT and RUN

CUT and RUN data were generated using the Epicypher Cutana CUT&RUN kit v3.0 (14-1048) according to the manufacturers provided instructions. Briefly 500k NALM6 cells per reaction were bound to 10μl of provided activated ConA beads in 0.2ml PCR tubes. Bead-bound cells were suspended in Antibody Buffer (Wash buffer with 0.1% digitonin, 0.5mM Spermidine, 2mM EDTA, and 1x HALT protease inhibitors) and incubated with 1ul PU.1 antibody (Cell Signaling 2258) or IgG (Epicypher 13-0042k) overnight on a nutator mixer at 4C. The next day after washing, pAG MNase was bound and targeted digestion was carried out for 2 hours at 4C. Digestion was stopped using 33μl Stop buffer + 1μl (0.5ng) E.coli spike-in DNA and then cleaved DNA were released for 10 minutes at 37C. DNA were then purified for library preparation using the included purification kit. >30M 75bp paired end reads were generated per sample using the Illumina Novaseq. The Nextflow CUT and RUN pipeline (v2.0) was used in spike-in mode to assess quality and map reads to the HG19 (human) and K12-MG1655 (E. coli) reference genomes (*73, 74*). The spike-in normalized .bam files from Nextflow were exported to Easeq (v1.111), where peaks were called against the IgG sample using adaptive local thresholding (p < 1×10^−5^, FDR < 1×10^−5^, Log_2_(Fold Change) > 1, merge within = 100bp, window size = 100bp) (*75*). Data shown are spike-in normalized bigwig files generated in Nextflow.

#### CRISPR/Cas9 deletion

rs1247117 deletions in Nalm6 were generated using CRISPR-Cas9 technology. In brief, one million Nalm6 cells were transiently transfected with precomplexed ribonuclear proteins (RNPs) consisting of 100pmol of each chemically modified sgRNA (Synthego, see **Sup File 9**), 35pmol of Cas9 protein (St. Jude Protein Production Core), and 3ug of ssODN (Alt-R modifications, IDT; see Table 1 below) via nucleofection (Lonza, 4D-Nucleofector™ X-unit) using solution P3 and program CV-104 in a large (100ul) cuvette according to the manufacturer’s recommended protocol. Three days post-nucleofection, genomic DNA was harvested via crude lysis and used for PCR amplification (see Sup File 3 for primers). The presence of the desired deletion was confirmed via gel electrophoresis and sequencing. To validate disruptions, targeted amplicons were generated using gene specific primers with partial Illumina adapter overhangs and sequenced. Cell pellets of approximately 10,000 cells were lysed and used to generate gene specific amplicons with partial Illumina adapters in PCR#1. Amplicons were indexed in PCR#2 and pooled with targeted amplicons from other loci to create sequence diversity. Additionally, 10% PhiX Sequencing Control V3 (Illumina) was added to the pooled amplicon library prior to running the sample on an Miseq Sequencer System (Illumina) to generate paired 2 × 250bp reads. Samples were demultiplexed using the index sequences, fastq files were generated, and NGS analysis was performed using CRIS.py (*76*).

### Statistical analysis

#### GWAS for *ex vivo* drug resistance in primary ALL cells

Multiple linear regression was used with log-transformed LC_50_ as dependent variable and genotype as independent variable. Genotypes were coded as 0, 1 and 2. Patient genetic ancestry was included in the linear model as covariates. Two tailed p-values were generated using a Wald test. All statistical analysis was performed in R v4.0.2. All SNVs with a p-value < 0.05 were further evaluated to determine if were eQTLs.

#### Gene expression profiling in primary ALL cells

Genotype information was correlated with RNA expression across patient samples to identify expression quantitative trait loci (eQTLs). eQTL mapping was performed using multiple linear regression and log-transformed FPKM gene expression as dependent variable and genotype as independent variable. Genotypes were coded as 0, 1 and 2. Patient genetic ancestry was included in the linear model as covariates. Two tailed p-values were generated using a Wald test. All statistical analysis was performed in R v4.0.2.

#### Promoter-capture Hi-C

Statistical modeling of chromatin looping was performed as described in the publication introducing the CHiCAGO tool (*71*). We used 5kb resolution and an adjusted P value cutoff of 0.05.

#### PU.1 binding affinity assay

A one-tailed student’s T-test was used to test the hypothesis that the alternate allele would show less affinity for PU.1 binding.

#### PU.1 CUT and RUN

PU.1 peaks were called against the IgG sample using adaptive local thresholding (p < 1×10^−5^, FDR < 1×10^−5^, Log_2_(Fold Change) > 1, merge within = 100bp, window size = 100bp) (*75*). Data shown are spike-in normalized bigwig files generated in Nextflow.

#### Nalm6 vincristine drug sensitivity assays

Individual Students T-tests were performed at each dose in each time point to test the hypothesis that deletion of the regulatory region containing rs1247117 in Nalm6 cells would alter sensitivity to vincristine.

#### Comparisons between groups of variant MPRA data

When comparing promoter-associated and distal promoter-connected variant MPRA data, the Mann-Whitney test was used. Comparisons within the distal promoter-connected variants from introns, UTRs and distal intergenic regions were carried out using the Kruskal-Wallis test with Dunn’s correction for multiple comparisons.

#### *CACUL1* and *EIF3A* qPCR and Dual Luciferase reporter assays

Student’s T tests were used to determine the significance of differences between samples.

## Supporting information

List of Supplemental Materials

## Data Availability

All non patient-associated data are available for reviewers on NCBI GEO GSE224204
Data from patient samples are available upon reasonable request via St. Jude cloud.

https://www.ncbi.nlm.nih.gov/geo/query/acc.cgi?acc=GSE224204

https://cloud.stjude.org

## Acknowledgments

We would like to thank the Hartwell Center at St. Jude for ATAC-seq, ChIP-seq and MPRA next-generation sequencing. We would also like to thank Jeremy Hunt and Brandon Smart for technical support. This work was performed at St. Jude Children’s Research Hospital and was also conducted in part by the ECOG-ACRIN Cancer Research Group. This work is supported by the National Cancer Institute (R01CA234490, P30CA021765, U10CA180820, UG1CA232760 and UG1CA189859), the National Institute of General Medical Studies (P50GM115279) and the American Lebanese Syrian Associated Charities (ALSAC). The content is solely the responsibility of the authors and does not necessarily represent the official views of the National Institutes of Health.

## Author contributions

Conceptualization: DS, RJM

Methodology: DS, KR Bhattarai, RJM

Investigation: KR Bhattarai, RJM, DCF, JDD, BPB

Formal Analysis: KR Bhattarai, RJM, KR Barnett, WY, CC

Data Curation: KR Barnett, WY, KRC, CC

Sample Acquisition: CSM, EJ, EP, MRL, SMK, WS, HI, SJ, CHP, MVR, JJY, WEE

Visualization: RJM, KR Bhattarai

Supervision: DS, RJM

Writing—original draft: DS, KR Bhattarai, RJM

Writing—review & editing: KR Bhattarai, RJM, KR Barnett, DCF, JDD, BPB, WY, KRC, CSM, EJ, EP, MRL, SMK, WS, HI, SJ, CHP, CC, MVR, JJY, WEE, DS

## Data and materials availability

Cell line ATAC-seq, Promoter capture HiC, RNA pol II, and PU.1 genomic binding data are available on GEO (GSE224204). Previously published H3K27Ac ChIP-seq data, “GSE175482_Nalm6_H3K27ac_0hr_merged.bw”, are located in GEO GSE175482. Patient sample and PDX associated genomics data are available upon request via St. Jude cloud (https://www.stjude.cloud/).

## Supplemental Materials

Supplemental Figures

Supplemental File 1 – SNP IDs, Sentinel SNVs, and associated Phenotypes of MPRA SNPs

Supplemental File 2 – MPRA source data

Supplemental File 3 – 556 reproducible and concordant regulatory variants

Supplemental File 4 – 53 reproducible and concordant regulatory variants at promotors or connected to promoters as determined by promoter CHiC.

Supplemental File 5 – Reference and alternative allele sequences used in dual-luciferase reporter assays

Supplemental File 6 – Nucleic acids associated with MPRA method

Supplemental File 7 – MPRA barcode sequences

Supplemental File 8 – PU.1 binding affinity assay probe sequences Supplemental File 9 – CRISPR/Cas9 rs1247117 deletion pool sequences

## SUPPLEMENTAL FIGURES

**Supplemental Figure 1.**
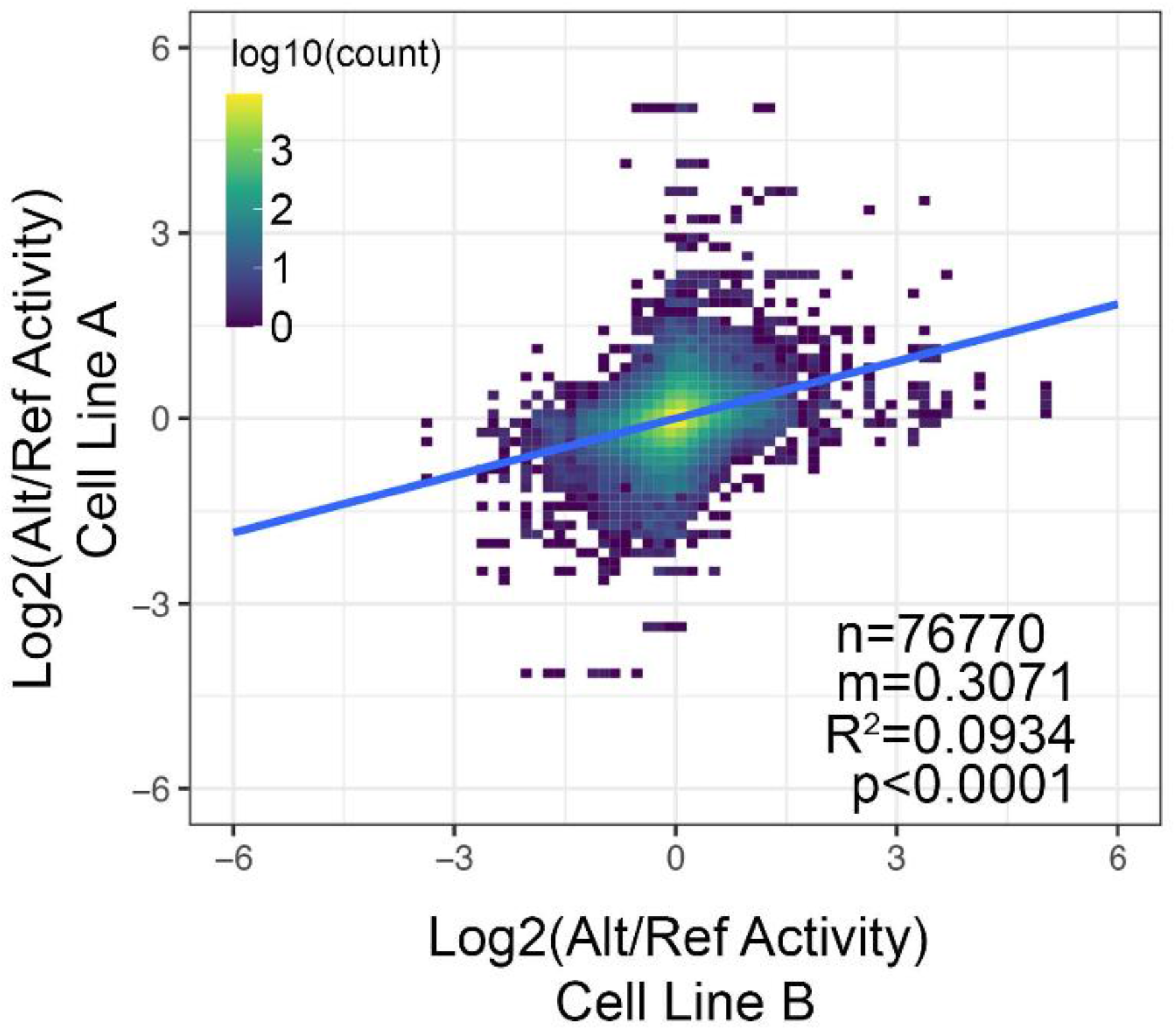
MPRA activity comparisons among all cell lines. Pair-wise linear correlation between changes in allele-specific transcriptional activity for all measurements and across all cell lines. R^2^ correlation and p-value are provided.

**Supplemental Figure 2.**
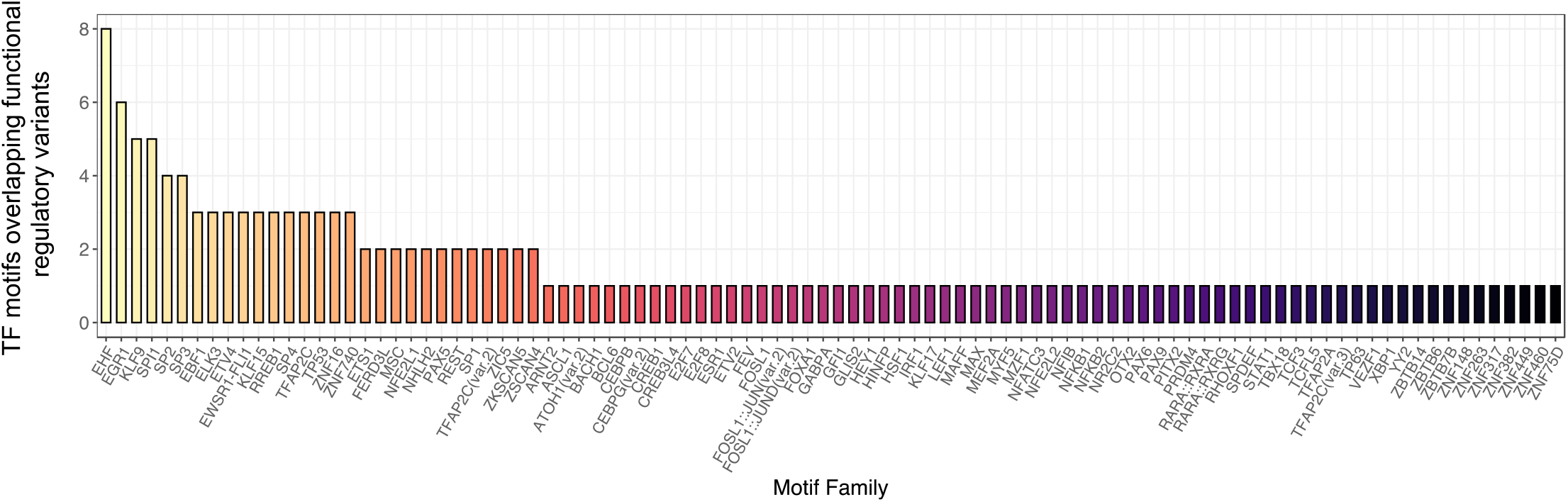
Transcription factor footprints at functional regulatory variants. Transcription factor (TF) footprints identified at 54 of 556 functional regulatory variants are shown and ranked by the total number of motifs identified.

**Supplemental Figure 3.**
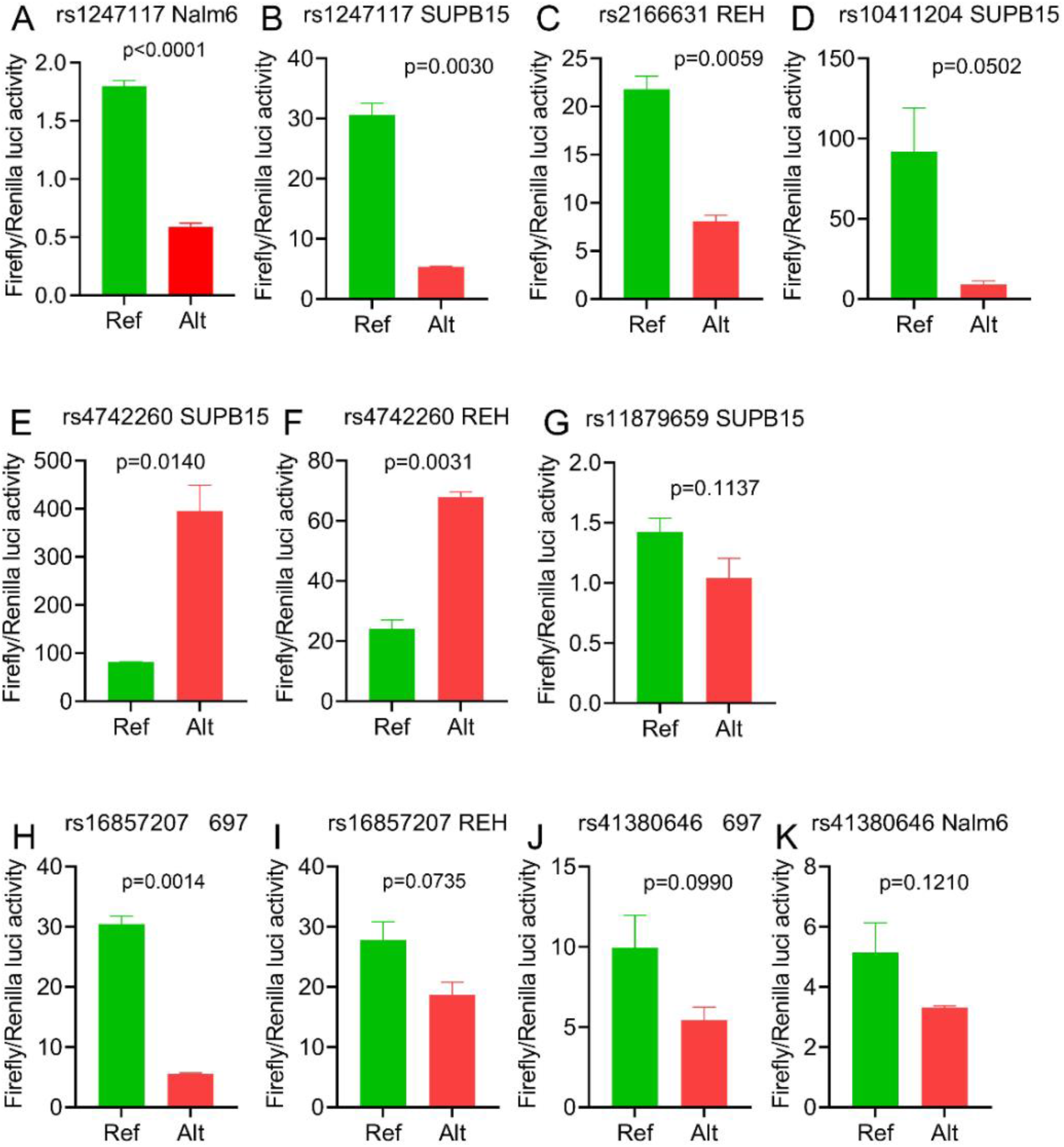
Dual-luciferase reporter assay validation of the indicated functional regulatory variant. **(A-K)** Dual-luciferase reporter assays comparing the reference (Ref, in green) and alternate (Alt, in red) alleles ability to drive luciferase expression is depicted. Variant rs number and the ALL cell line the luciferase reporter assay was tested in is provided. Data show the mean +/- SEM of three **(A)** or two **(B-K)** independent experiments. P-value is calculated using a student’s t-test.

**Supplemental Figure 4.**
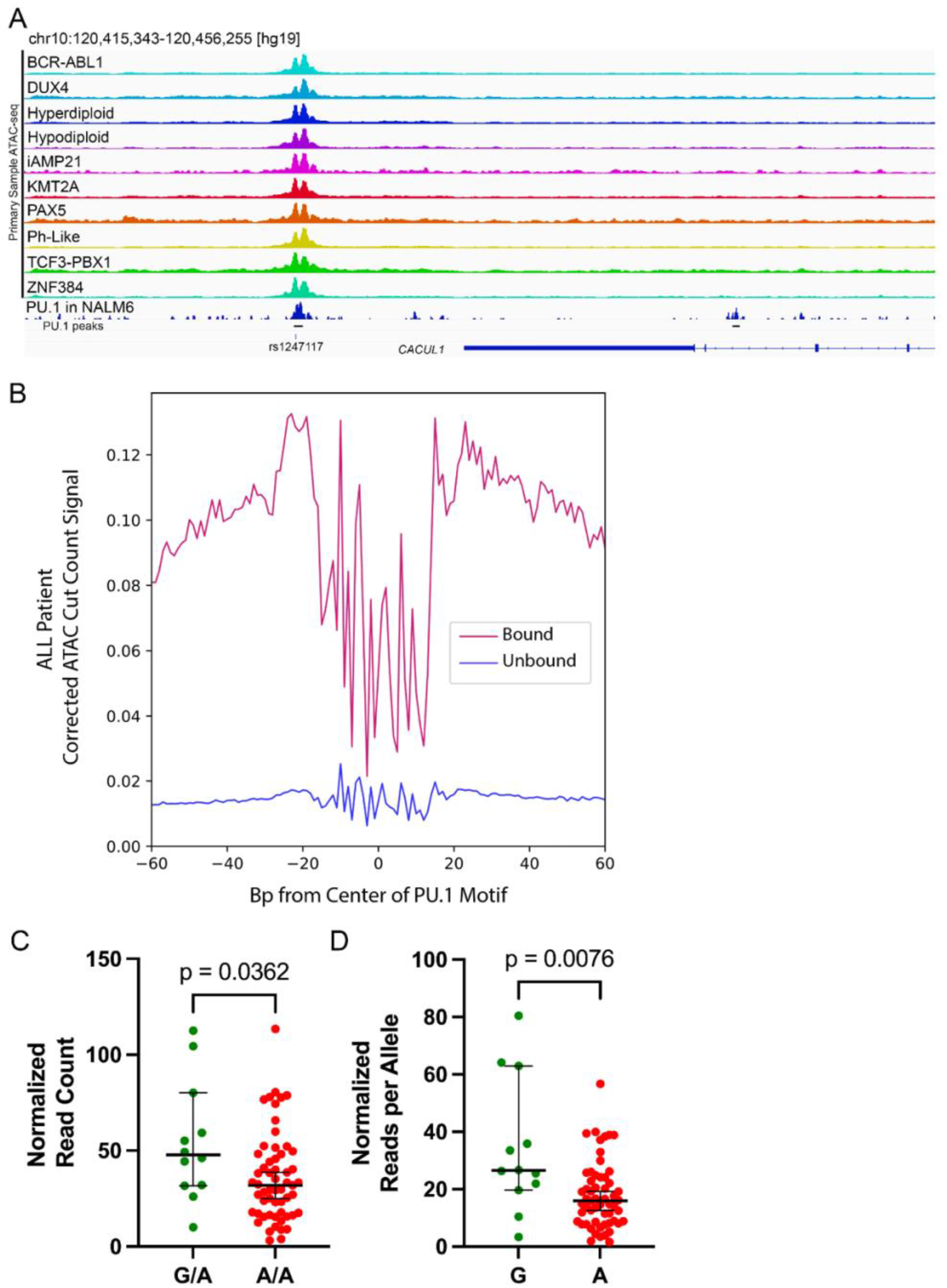
Chromatin accessibility at rs1247117 in primary ALL cells. **(A)** IGV genome browser image of ATAC-seq chromatin accessibility spanning rs1247117 in diverse molecular subtypes of ALL is provided. **(B)** PU.1 footprint analysis comparing normalized ATAC-seq cut count signal for all bound PU.1 sites (red) compared to unbound (blue) sites across all primary ALL cells from patients. **(C)** Primary ALL cells with SNV genotype information were analyzed (n=69). Normalized ATAC-seq read counts in heterozygous (GA) primary ALL cells (n=12) at rs1247117 compared to homozygous (AA) primary ALL cells (n=57). Mann Whitney U test p-value is provided. **(D)** Normalized ATAC-seq read counts per allele in primary ALL cells for G allele (n=12) and A allele (n=69). Normalized counts for G and A alleles are shown. Mann Whitney U test p-value is provided.

