## Supplemental File 5 for "Functional investigation of inherited noncoding genetic variation impacting the pharmacogenomics of childhood acute lymphoblastic leukemia treatment"

**Reference and alternative allele sequences used in dual-luciferase reporter assays**

1. **snpID:** rs1247117

**Alleles:** G/A

**Position (original):** chr10:120428802

**Altered allele (-150/+150bp):** chr10:120428652-120428952

**Reference Sequence**

CTGCCCATCAGAGCCCATTAGACCAGAATGAGACTCCCAGGGTGGCCGCAGAGCAGCAGCATTTCAGCTCATCAGAGGAAGCTGCCAAATGAGCCAGTCTGAGCAGCATTTGGGGGTAGATGTTCGCTTCCTATTAACAGGCTAAATTGCGGAAGTGGCTCAGAGTTTCATTCCCACTGTGCCACTCCCTGGGGTCTGCAGCGTGGCCTGGGAGAAAAAGGCACAGCCTCGGGGTCAGACCCGAGTTGGACCCAGCTCTGTCCCCTGGTACCTGAGGGACTCTGGATAAATTACTGGATTT

**Reverse Strand: Ref Allele (C)**

AAATCCAGTAATTTATCCAGAGTCCCTCAGGTACCAGGGGACAGAGCTGGGTCCAACTCGGGTCTGACCCCGAGGCTGTGCCTTTTTCTCCCAGGCCACGCTGCAGACCCCAGGGAGTGGCACAGTGGGAATGAAACTCTGAGCCACTTCCGCAATTTAGCCTGTTAATAGGAAGCGAACATCTACCCCCAAATGCTGCTCAGACTGGCTCATTTGGCAGCTTCCTCTGATGAGCTGAAATGCTGCTGCTCTGCGGCCACCCTGGGAGTCTCATTCTGGTCTAATGGGCTCTGATGGGCAG

**Reverse Strand: Alt allele (T)**

AAATCCAGTAATTTATCCAGAGTCCCTCAGGTACCAGGGGACAGAGCTGGGTCCAACTCGGGTCTGACCCCGAGGCTGTGCCTTTTTCTCCCAGGCCACGCTGCAGACCCCAGGGAGTGGCACAGTGGGAATGAAACTCTGAGCCACTTCTGCAATTTAGCCTGTTAATAGGAAGCGAACATCTACCCCCAAATGCTGCTCAGACTGGCTCATTTGGCAGCTTCCTCTGATGAGCTGAAATGCTGCTGCTCTGCGGCCACCCTGGGAGTCTCATTCTGGTCTAATGGGCTCTGATGGGCAG

2. **snpID:** rs10411204

**Alleles:** A/G

**Position (original):** chr19:50507336

**Altered allele (-150/+150bp):** chr19:50507186-50507486

**Reference Sequence**

CAGTGCTCACGCCTATAATCCCAACACTTTGGGAGGCAGAGGCAGGAGGATGGCTTGAGGCCAAGAGTTTGAGACCAGCCTGGCCAACATAGTGAGATCGTCTCTACTTTAAAAAATAAATAAATAAAGGGACTGCTGAAGGAATGGATGACTCACACAGGCTGGGCACTGGGGTTTGCTATTTTTATCAAACAACCTGAGTCATATAAGGGAAAACCAAAACCACATTGGCCCAGAGTAGCTCCTCCTGTGCTTCCACTGTGGGTTTCTGGGGGAGGTCAAGCGTGGTTTCTTCTCTCAG

**Reverse Strand: Ref Allele (T)**

CTGAGAGAAGAAACCACGCTTGACCTCCCCCAGAAACCCACAGTGGAAGCACAGGAGGAGCTACTCTGGGCCAATGTGGTTTTGGTTTTCCCTTATATGACTCAGGTTGTTTGATAAAAATAGCAAACCCCAGTGCCCAGCCTGTGTGAGTCATCCATTCCTTCAGCAGTCCCTTTATTTATTTATTTTTTAAAGTAGAGACGATCTCACTATGTTGGCCAGGCTGGTCTCAAACTCTTGGCCTCAAGCCATCCTCCTGCCTCTGCCTCCCAAAGTGTTGGGATTATAGGCGTGAGCACTG

**Reverse Strand: Alt allele (C)**

CTGAGAGAAGAAACCACGCTTGACCTCCCCCAGAAACCCACAGTGGAAGCACAGGAGGAGCTACTCTGGGCCAATGTGGTTTTGGTTTTCCCTTATATGACTCAGGTTGTTTGATAAAAATAGCAAACCCCAGTGCCCAGCCTGTGTGAGCCATCCATTCCTTCAGCAGTCCCTTTATTTATTTATTTTTTAAAGTAGAGACGATCTCACTATGTTGGCCAGGCTGGTCTCAAACTCTTGGCCTCAAGCCATCCTCCTGCCTCTGCCTCCCAAAGTGTTGGGATTATAGGCGTGAGCACTG

3. **snpID:** rs11879659

**Alleles:** C/T

**Position (original):** [chr19:50490522](https://genome.ucsc.edu/cgi-bin/hgTracks?hgsid=1346024607_9rXZtzzHLLLQxEYym69vmEaoh98G&db=hg19&position=chr19%3A50490522-50490522)

**Altered allele (-150/+150bp):** chr19:50490372-50490672

**Reference Sequence**

CCATCTGCCTTGGCCTCCCAAAGTGCTGGGATTAAGGCGTGAACCACCGTGCCCGGCCTGGGCAACTGATATTCATGTCTGAAGTGCATGCCTAGCACGCCCGCTAGGATCAGCTGCCCACTGTCCTTGGTGCTGAACTCATACCCACAGCGCTTCCCTTTTGTGCTCCGCTGTCCCTGGTGCTGAACCCACATCCCTAGCACTTCCCTTTTGTGTCCCGCTGCCTGCTGTTCTTGGTGCTGAACACACACCTTAGTGCCTCCCTTTTGTGCCCAGCTGCCTGCTGTCCCTAGTGCTGAAC

**Reverse Strand: Ref Allele (G)**

GTTCAGCACTAGGGACAGCAGGCAGCTGGGCACAAAAGGGAGGCACTAAGGTGTGTGTTCAGCACCAAGAACAGCAGGCAGCGGGACACAAAAGGGAAGTGCTAGGGATGTGGGTTCAGCACCAGGGACAGCGGAGCACAAAAGGGAAGCGCTGTGGGTATGAGTTCAGCACCAAGGACAGTGGGCAGCTGATCCTAGCGGGCGTGCTAGGCATGCACTTCAGACATGAATATCAGTTGCCCAGGCCGGGCACGGTGGTTCACGCCTTAATCCCAGCACTTTGGGAGGCCAAGGCAGATGG

**Reverse Strand: Alt allele (A)**

GTTCAGCACTAGGGACAGCAGGCAGCTGGGCACAAAAGGGAGGCACTAAGGTGTGTGTTCAGCACCAAGAACAGCAGGCAGCGGGACACAAAAGGGAAGTGCTAGGGATGTGGGTTCAGCACCAGGGACAGCGGAGCACAAAAGGGAAGCACTGTGGGTATGAGTTCAGCACCAAGGACAGTGGGCAGCTGATCCTAGCGGGCGTGCTAGGCATGCACTTCAGACATGAATATCAGTTGCCCAGGCCGGGCACGGTGGTTCACGCCTTAATCCCAGCACTTTGGGAGGCCAAGGCAGATGG

4. **snpID:** rs12660691

**Alleles:** A/C

**Position (original):**  chr6:130008445

**Altered allele (-150/+150bp):** chr6:130008295-130008595

**Reference Sequence**

TTCATCCCTTTGAGATGACAGTTGGCTACAGATGTTGAACAGGTATGCAGGGACCAGTTTCTATGCACCCCACCTTTAGGACTTGCAACTCTAGTTCACGGGTCTCAGAGAAGTTGTAAGTTACCAGTTTCCAACATTCCAAATTACTGCAAATGCCAGAAACTACAAATGTTTGGAAATGCCAAGAGTCTGAGGATCTACTGACACAGCATATAGCTGAAACCTCACCCACATAGTGTTAAGAAAGCGTTTCCTGCCTGTCTGGGGCTGACAAACCACAACAACCCTTTTCTTCTTCCTC

**Reverse Strand: Ref Allele (T)**

GAGGAAGAAGAAAAGGGTTGTTGTGGTTTGTCAGCCCCAGACAGGCAGGAAACGCTTTCTTAACACTATGTGGGTGAGGTTTCAGCTATATGCTGTGTCAGTAGATCCTCAGACTCTTGGCATTTCCAAACATTTGTAGTTTCTGGCATTTGCAGTAATTTGGAATGTTGGAAACTGGTAACTTACAACTTCTCTGAGACCCGTGAACTAGAGTTGCAAGTCCTAAAGGTGGGGTGCATAGAAACTGGTCCCTGCATACCTGTTCAACATCTGTAGCCAACTGTCATCTCAAAGGGATGAA

**Reverse Strand: Alt allele (G)**

GAGGAAGAAGAAAAGGGTTGTTGTGGTTTGTCAGCCCCAGACAGGCAGGAAACGCTTTCTTAACACTATGTGGGTGAGGTTTCAGCTATATGCTGTGTCAGTAGATCCTCAGACTCTTGGCATTTCCAAACATTTGTAGTTTCTGGCATTGGCAGTAATTTGGAATGTTGGAAACTGGTAACTTACAACTTCTCTGAGACCCGTGAACTAGAGTTGCAAGTCCTAAAGGTGGGGTGCATAGAAACTGGTCCCTGCATACCTGTTCAACATCTGTAGCCAACTGTCATCTCAAAGGGATGAA

5. **snpID:** rs41380646

**Alleles:** C/T

**Position (original):** chr15:64185530

**Altered allele (-150/+150bp):** chr15:64185380-64185680

**Reference Sequence**

CAGAGTGGTTGCCAGGGGCAGAACCACTATAGGGGATCTTCCTTCCTCTTTTGCTCCAGGCTCCTCACTTACATGTCTATTTTTACCCTGTACTGTATGCTGTCAATAAGAAGCATGGTTATTACAGCAGCAACCACATTCCGAGTGCAACGGAACGGGAACTTCACATGCATCAGCTCATTCCATTTCACCCTCACAGAAGCCCTGCGGGAGCATACTGTCATTCCCATTTTACAAGAGGGAGAACTGAGGCTCAGTTCGTTGCTAACAGTTACCCAGCCAAGAAGGTGCCAGAACTGGG

**Reverse Strand: Ref Allele (G)**

CCCAGTTCTGGCACCTTCTTGGCTGGGTAACTGTTAGCAACGAACTGAGCCTCAGTTCTCCCTCTTGTAAAATGGGAATGACAGTATGCTCCCGCAGGGCTTCTGTGAGGGTGAAATGGAATGAGCTGATGCATGTGAAGTTCCCGTTCCGTTGCACTCGGAATGTGGTTGCTGCTGTAATAACCATGCTTCTTATTGACAGCATACAGTACAGGGTAAAAATAGACATGTAAGTGAGGAGCCTGGAGCAAAAGAGGAAGGAAGATCCCCTATAGTGGTTCTGCCCCTGGCAACCACTCTG

**Reverse Strand: Alt Allele (A)**

CCCAGTTCTGGCACCTTCTTGGCTGGGTAACTGTTAGCAACGAACTGAGCCTCAGTTCTCCCTCTTGTAAAATGGGAATGACAGTATGCTCCCGCAGGGCTTCTGTGAGGGTGAAATGGAATGAGCTGATGCATGTGAAGTTCCCGTTCCATTGCACTCGGAATGTGGTTGCTGCTGTAATAACCATGCTTCTTATTGACAGCATACAGTACAGGGTAAAAATAGACATGTAAGTGAGGAGCCTGGAGCAAAAGAGGAAGGAAGATCCCCTATAGTGGTTCTGCCCCTGGCAACCACTCTG

6. **snpID:** rs16857207

**Alleles:** G/A

**Position (original):** chr3:182967025

**Altered allele (-150/+150bp):** chr3:182966875-182967175

**Reference Sequence: Ref allele (G)**

TCTTTTAAAATATGTGTTAATCAGCACTCAAGATTGTTCCTACAAAAATGTCACTCCCTCACTCAATTTGCATGTGCCCTCCTGAGTAGGAGAGAAAGAGCTGAGTTAGAGGCAGCCCTCTGGGACCTGCACAGAACCCTGTATGCTTCCGGGAGTGTGGAGTGTGTGTCTGATCTGCTGCTGGGAAAAGGAGAAGAAATGCTGAGACACTCACTGCCAGGGGCTGGTATCAGGCCATTTTCACAGGGGCTGTTGGAGGGTTGACAGCACAGCTCTACTGGACCAGGGAGGGTCCCAGCCA

**Forward strand: Alt allele (A)**

TCTTTTAAAATATGTGTTAATCAGCACTCAAGATTGTTCCTACAAAAATGTCACTCCCTCACTCAATTTGCATGTGCCCTCCTGAGTAGGAGAGAAAGAGCTGAGTTAGAGGCAGCCCTCTGGGACCTGCACAGAACCCTGTATGCTTCCAGGAGTGTGGAGTGTGTGTCTGATCTGCTGCTGGGAAAAGGAGAAGAAATGCTGAGACACTCACTGCCAGGGGCTGGTATCAGGCCATTTTCACAGGGGCTGTTGGAGGGTTGACAGCACAGCTCTACTGGACCAGGGAGGGTCCCAGCCA

7. **snpID:** rs2166631

**Alleles:** C/T

**Position (original):** chr10:73472882

**Altered allele (-150/+150bp):** chr10:73472732-73473032

**Reference Sequence: Ref allele (C)**

CCCTCTCCCATCCCAGGGAGTGTGGGCCCCCAGGTTGCAGAGCTGAGGATAGGGCTCTGGCTCCCCACAAACAGGAGGAAGACTTGGCCTCCCCCTGCATATCTCCCGCCCCACCCAGAAGGGCCTTTCAGAGTAGCGGGGAGAAAGAGACGTCACAATTTCCCGGAAAGGAGTTGAGAAGAGAGCCAGGGCCCAGCAAGGCCCAGAACAGAGCTGCTTTCACAGAGCCCTTGGCCGAGGCTCCCCTCTTGGGAAGTAAACAGGCACAAGAAGAAAGGGCTCCTTTGCCCAGGGAGCAGGC

**Forward strand: Alt allele (T)**

CCCTCTCCCATCCCAGGGAGTGTGGGCCCCCAGGTTGCAGAGCTGAGGATAGGGCTCTGGCTCCCCACAAACAGGAGGAAGACTTGGCCTCCCCCTGCATATCTCCCGCCCCACCCAGAAGGGCCTTTCAGAGTAGCGGGGAGAAAGAGATGTCACAATTTCCCGGAAAGGAGTTGAGAAGAGAGCCAGGGCCCAGCAAGGCCCAGAACAGAGCTGCTTTCACAGAGCCCTTGGCCGAGGCTCCCCTCTTGGGAAGTAAACAGGCACAAGAAGAAAGGGCTCCTTTGCCCAGGGAGCAGGC

8. **snpID:** rs4742260

**Alleles:** T/C

**Position (original):** chr9:6757031-6757031

**Altered allele (-150/+150bp):** chr9:6756881-6757181

**Reference Sequence: Ref allele (T)**

CGATATATTTTTAAAATTCTGTTATCAGTATAATGTAACAGAAAATTCAGCATATTAGCCCAACTTTGCTCACTGGTGTGAAGATATGCCCAATCTTTCTTAGCTAAGACGTTAGAGAAAACGCCACAAGGGACCCAGCTGGGAGCTCTGTAGGAAGTTTAAAAAAAAAATCATTCAGCTTTGCAAAACATCTGATGCTCAACCATCTGTCTTTGGCAGTACAGGGGCTTAAGGTCTGGTTGGTTCCTTCTGACACCTCGGAAGGCGGAGTTACAGAGGAAAAGCAACAAGTTGTAGGAAG

**Forward strand: Alt allele (C)**

CGATATATTTTTAAAATTCTGTTATCAGTATAATGTAACAGAAAATTCAGCATATTAGCCCAACTTTGCTCACTGGTGTGAAGATATGCCCAATCTTTCTTAGCTAAGACGTTAGAGAAAACGCCACAAGGGACCCAGCTGGGAGCTCTGCAGGAAGTTTAAAAAAAAAATCATTCAGCTTTGCAAAACATCTGATGCTCAACCATCTGTCTTTGGCAGTACAGGGGCTTAAGGTCTGGTTGGTTCCTTCTGACACCTCGGAAGGCGGAGTTACAGAGGAAAAGCAACAAGTTGTAGGAAG
